# Antimicrobial susceptibility patterns of respiratory Gram-negative bacterial isolates from COVID-19 patients in Switzerland

**DOI:** 10.1101/2021.03.10.21253079

**Authors:** Marina Gysin, Claudio Tirso Acevedo, Klara Haldimann, Elias Bodendoerfer, Frank Imkamp, Karl Bulut, Philipp Karl Buehler, Silvio Daniel Brugger, Katja Becker, Sven N. Hobbie

## Abstract

**Background:** Bacterial superinfections associated with COVID-19 are common in ventilated ICU patients and impact morbidity and lethality. However, the contribution of antimicrobial resistance to the manifestation of bacterial infections in these patients has yet to be elucidated.

**Methods:** We collected 70 Gram-negative bacterial strains, isolated from the lower respiratory tract of ventilated COVID-19 patients in Zurich, Switzerland between March and May 2020. Species identification was performed using MALDI-TOF; antibiotic susceptibility profiles were determined by EUCAST disk diffusion and CLSI broth microdilution assays. Selected *Pseudomonas aeruginosa* isolates were analyzed by whole-genome sequencing.

**Results:** *P. aeruginosa* (46%) and *Enterobacterales* (36%) comprised the two largest etiologic groups. Drug resistance in *P. aeruginosa* isolates was high for piperacillin/tazobactam (65.6%), cefepime (56.3%), ceftazidime (46.9%) and meropenem (50.0%). *Enterobacterales* isolates showed slightly lower levels of resistance to piperacillin/tazobactam (32%), ceftriaxone (32%), and ceftazidime (36%). All *P. aeruginosa* isolates and 92% of *Enterobacterales* isolates were susceptible to aminoglycosides, with apramycin found to provide best-in-class coverage. Genotypic analysis of consecutive *P. aeruginosa* isolates in one patient revealed a frameshift mutation in the transcriptional regulator *nalC* that coincided with a phenotypic shift in susceptibility to β-lactams and quinolones.

**Conclusions:** Considerable levels of antimicrobial resistance may have contributed to the manifestation of bacterial superinfections in ventilated COVID-19 patients, and may in some cases mandate consecutive adaptation of antibiotic therapy. High susceptibility to amikacin and apramycin suggests that aminoglycosides may remain an effective second-line treatment of ventilator-associated bacterial pneumonia, provided efficacious drug exposure in lungs can be achieved.

## BACKGROUND

The emergence of the coronavirus disease 2019 (COVID-19) has resulted in high rates of intensive care unit (ICU) admissions of critically ill patients [1] suffering from acute respiratory distress syndrome (ARDS) [2,3]. Respiratory viral infections predispose patients to secondary bacterial infections which are associated with increased morbidity and case fatality rates [4,5]. Particularly, secondary bacterial infections acquired in the setting of ICU-treatment are also independently associated with higher risk of mortality when compared with community-acquired infections [6].

Secondary bacterial infections in COVID-19 patients, hereafter referred to as superinfections, have not been intensively investigated thus far. Yet, mainly small cohort studies have reported high superinfection rates in critically ill and/or deceased COVID-19 patients. For instance, a retrospective cohort study by Zhou *et al*. documented secondary infections in 50% of deceased COVID-19 patients and ventilator-associated pneumonia (VAP) in a third of mechanically ventilated patients, despite the fact that 95% of patients received antibiotic treatment [7]. A study by Du *et al*. described secondary bacterial infections at a late stage of disease in 10 of 21 deceased patients with the etiological spectrum including *Klebsiella pneumoniae, Staphylococcus spp*., *Acinetobacter baumannii* and *Escherichia coli* [8]. However, a comprehensive antimicrobial resistance (AMR) surveillance of bacterial superinfection in critically ill COVID-19 patients treated in the ICU has yet to be reported.

In recent years, the incidence of infections caused by gram-negative bacilli (GNB), especially multidrug-resistant (MDR) organisms, in highly susceptible ICU patients has increased [9,10]. Particularly infections caused by MDR GNB are associated with a substantial risk of morbidity and in-hospital mortality for the critically ill [6]. The increased exposure of patients to antimicrobials and the global surge in hospital and especially ICU admissions due to the COVID-19 pandemic are rising concerns on the long-term impact on AMR in the acute care setting [11–13]. During the beginning of the COVID-19 pandemic, uncertainty regarding the novel disease has resulted in high antibiotic use since empirical antibiotic treatment was commonly prescribed before or at the time of hospital admission. A meta-analysis by Rawson *et al*. revealed that 72% of hospitalized patients received antimicrobial therapy despite low initial evidence of community-acquired co-infections at that time [14]. Similarly, the ISARIC WHO Clinical Characterization Protocol reported preceding anti-infective treatment in 76.6% of hospitalized patients [15].

A detailed understanding of the epidemiology and AMR pattern of bacterial superinfections in critically ill COVID-19 patients is urgently needed for adequate treatment. Here we report the bacterial spectrum and the antimicrobial susceptibility patterns of respiratory GNB isolated from ventilated ICU patients during the first COVID-19 wave in Switzerland. In addition, we investigate changes in the antimicrobial resistance over time for a single *P. aeruginosa* infection under antibiotic therapy using whole-genome sequencing (WGS).

## METHODS

### Study population

This study was conducted within the MicrobiotaCOVID cohort, a single-center, prospective observational study conducted at the University Hospital Zurich, Switzerland. The study was approved by the Local Ethics Committee of the Canton of Zurich, Switzerland (Kantonale Ethikkommission Zurich BASEC ID 2020 – 00646). All patients with confirmed SARS-CoV-2 infection admitted to the ICU requiring mechanical ventilation between March and May 2020 were included (*n* = 40). SARS-CoV-2 was detected by real-time RT-PCR as previously described [16]. Informed consent of all patients was obtained. The study cohort during this time period has been described recently [17].

### Sample collection

Longitudinal sample collection, processing and testing was performed as described recently [17]. In brief, the following sampling was performed: If the clinical situation allowed, bronchoalveolar lavages (BAL) with 10ml of saline were collected by the ICU personnel upon ICU admission and during the later course of the disease if clinically indicated. Tracheobronchial secretions (TBS) were collected from each intubated patient at least on day 0 (i.e., upon ICU admission), day 1, day 2, day 3, day 5 and henceforth every 5 days. If the clinical situation as determined by the ICU attending physician did not allow TBS collection, no sampling was performed. Samples were initially processed at the diagnostic laboratory of the Institute of Medical Microbiology (IMM) in the course of routine diagnostics. Species identification was performed with MALDI-TOF MS (Bruker Daltonics, Bremen, Germany) using the direct formic acid transfer method [18]. Repetitive detected isolates of the same species in the same patients were included to monitor changes in antimicrobial susceptibility over time.

### Antimicrobial susceptibility testing (AST)

Bacterial isolates were inoculated onto Columbia sheep blood (COS) agar and were incubated for 24 hours prior to antimicrobial susceptibility testing. The European Committee on Antimicrobial Susceptibility Testing (EUCAST) disk diffusion method (version 8.0, January 2020) [19] was applied to determine the isolate’s antimicrobial susceptibility to piperacillin/tazobactam (TPZ36), amoxicillin/clavulanate (AMC30), ceftriaxone (CRO30), cefepime (FEP30), meropenem (MEM10), amikacin (AK30), tobramycin (TOB10), gentamicin (CN10), trimethoprim/sulfamethoxazole (SXT25) ceftazidime (CAZ10) and ciprofloxacin (CIP5). The antibiotic SirScan Disks were obtained from i2a Diagnostics, Montpellier, France. Interpretative criteria in the EUCAST guidelines 2020 [20] were applied to set clinical resistance breakpoints and translate inhibition zone diameters into either resistant (R) or non-resistant (susceptible, S and “susceptible, increased exposure” (intermediate), I) phenotypes. Interpretative criteria for *P. aeruginosa* resistance to GEN were derived from the EUCAST guidelines 2019 [21] instead, since they have been removed from the 2020 version. Interpretative criteria for *Burkholderia cenocepacia* resistance to MEM and SXT (Table S4) were derived from the Clinical and Laboratory Standards Institute (CLSI) guidelines 2020 [22].

Antimicrobial susceptibilities were further assessed by broth microdilution assays following the CLSI guidelines [22,23]. Interpretative criteria in the EUCAST guidelines 2020 were followed to set clinical breakpoints and translate MICs into drug susceptibility. Interpretative criteria for *P. aeruginosa* resistance to GEN were derived from the EUCAST guidelines 2019 instead, since they have been removed from the 2020 version. For the aminoglycoside plazomicin, the FDA-identified interpretative criteria of 2, 4, and ≥8 mg/L were used for susceptible, intermediate, and resistant, respectively [24].

The minimal inhibitory concentration (MIC) performance standard for the aminoglycoside apramycin was set to a modal value of 4 mg/L for *E. coli* ATCC 25922 and an acceptable range of 2-8 mg/L. Epidemiological cut-off values (ECOFFs) of 8 mg/L for *Enterobacterales* and 16 mg/L for *P. aeruginosa* were used as interpretative criteria [25].

### Whole-Genome Sequencing of selected *P. aeruginosa* isolates

Whole-genome sequencing was applied to detect resistance determinants of eight selected consecutive *P. aeruginosa* isolates from a single patient.

DNA was extracted from cultures of the clinical isolates grown on Columbia sheep blood (COS) agar plates using the DNeasy® UltraClean® Microbial kit (Qiagen, Hilden, Germany) according to the manufacturer’s recommendations. Library preparation was performed using the Qiagen® QIAseq FX DNA kit (Qiagen, Hilden, Germany), according to manufacturer’s instructions. Sequencing library quality and size distribution were analyzed on a fragment analyzer automated CE system (Advanced Analytical Technologies Inc., Heidelberg, Germany), using the fragment analyzer 474 HS next generation sequencing (NGS) kit. Sequencing libraries were pooled in equimolar concentrations and paired-end sequenced (2 × 150 bp) on an Illumina MiSeq platform (Illumina®, San Diego, CA, USA).

Raw sequencing reads (FASTQ) were filtered and trimmed using Trimmomatic [26]. CONTIGS were assembled from processed FASTQ files using SPAdes (v3.13.0). To identify antibiotic resistance determinants CONTIGS were analyzed using the command line version of Resistance Gene Identifier (RGI; v4.2.2) and CARD (Comprehensive Antibiotic Resistance Database, v3.1.0) [27,28].

## RESULTS

### Species distribution

Out of a total of 314 respiratory samples (289 TBS and 25 BAL) 168 GNB isolates were detected, in 19 of the 40 patients (48%). A representative subset of 70 GNB, including repetitive isolates from 17 patients, were further analyzed in this study. More information about the 17 patients is provided in the Supplementary Information (Table S1).

The two largest groups of pathogens in our panel of COVID-19 GNB isolates were *Pseudomonas aeruginosa* (46%) and *Enterobacterales* (36%). *Burkholderia cenocepacia* (17%) was found in smaller numbers, as well as a single *Acinetobacter bereziniae* isolate (Figure 1A). Within the order of *Enterobacterales, Enterobacter cloacae* (32%) and *Klebsiella pneumoniae* (28%) were the two predominant species, followed by *Klebsiella aerogenes* (20%), *Citrobacter spp*. (16%), and *Escherichia coli* (4%) (Figure 1B). The species distribution of the analyzed subset (*n* = 70) was shown to be representative for all the identified Gram-negative isolates of the MicrobiotaCOVID cohort (*n* = 168, Figure S1).

**Figure 1.**
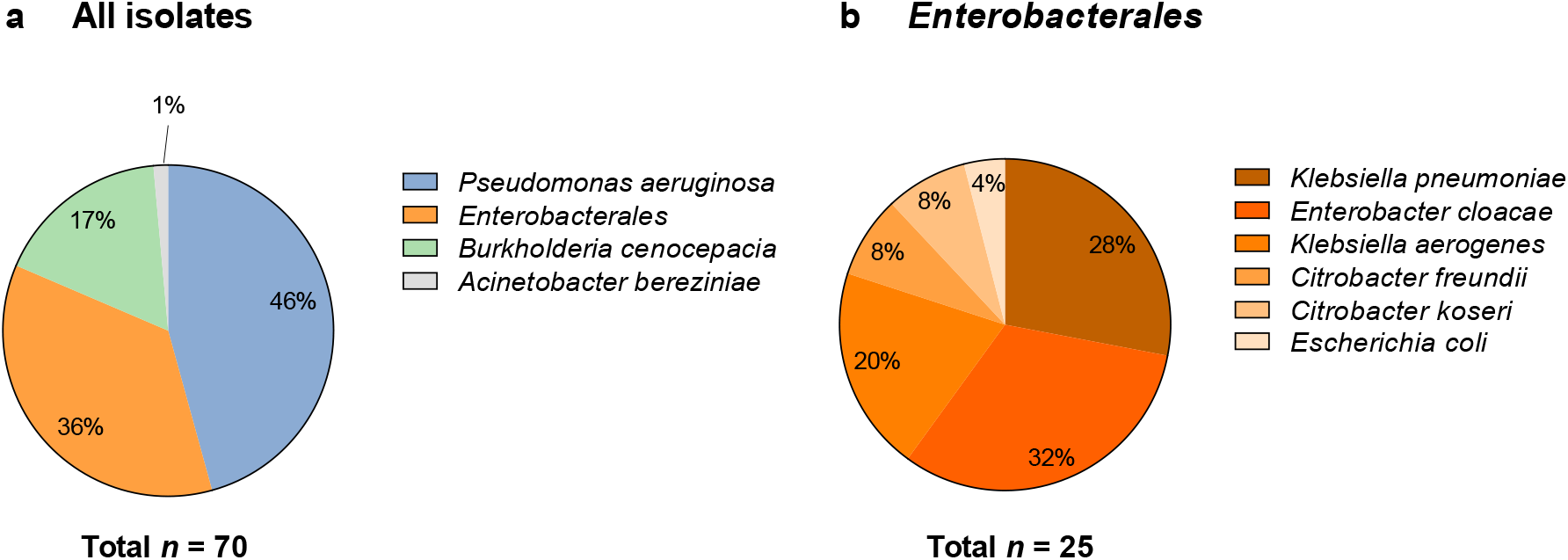
Subset distribution of all Gram-negative respiratory isolates studied (a) and further differentiation within the order of *Enterobacterales* (b).

### Antimicrobial susceptibility patterns

The antimicrobial susceptibility profile of all *Enterobacterales* and *P. aeruginosa* isolates (*n* = 57) was determined by the EUCAST disk diffusion method (Figure 2, Table S2).

**Figure 2.**
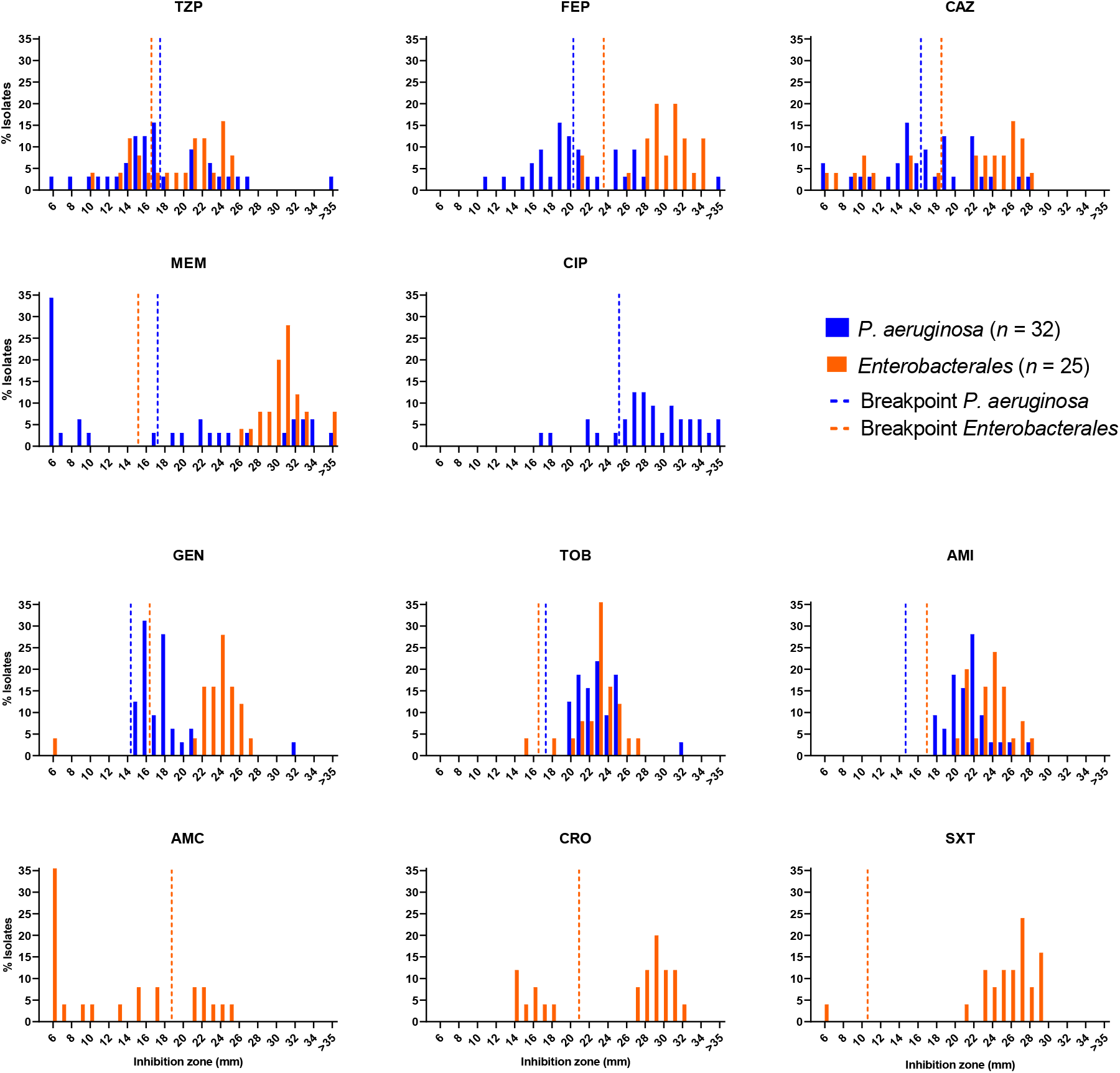
Inhibition zone diameter distributions for respiratory *Enterobacterales* and *P. aeruginosa* isolates from ventilated COVID-19 patients. Vertical dashed lines indicate the EUCAST clinical resistant breakpoint values for piperacillin/tazobactam (TZP), cefepime (FEP), ceftazidime (CAZ), meropenem (MEM), ciprofloxacin (CIP), gentamicin (GEN), tobramycin (TOB), amikacin (AMI), amoxicillin/clavulanate (AMC), ceftriaxone (CRO) and trimethoprim/sulfamethoxazole (SXT) for *Enterobacterales* (orange) and *P. aeruginosa* (blue) isolates, respectively.

A high proportion of *P. aeruginosa* isolates was found to be resistant to the standard-of-care antibiotics cefepime (FEP, 56.3%), ceftazidime (CAZ, 46.9%), and meropenem (MEM, 50.0%). *P. aeruginosa* resistance to piperacillin/tazobactam (TZP, 65.6%) was the highest for any of the relevant drugs tested in this study. Resistance to ciprofloxacin (CIP) was comparatively low in *P. aeruginosa* isolates (15.6%).

*Enterobacterales* isolates showed likewise resistance to TZP (32.0%), ceftriaxone (CRO, 32.0%), and CAZ (36.0%). The *Enterobacterales* resistance was lower for FEP (8.0%) and trimethoprim/sulfamethoxazole (SXT, 4.0%).

Disk diffusion further revealed all the *P. aeruginosa* isolates to be susceptible to gentamicin (GEN), tobramycin (TOB), and amikacin (AMI). Aminoglycoside susceptibility was also high for the *Enterobacterales* isolates with only a single *E. coli* isolate lacking susceptibility to gentamicin and tobramycin.

To study the antimicrobial susceptibility profiles in more detail and to test additional antibiotics not routinely included in the disk diffusion panel, the clinical isolates were also analyzed by broth microdilution assays to determine the minimal inhibitory concentration (MIC) (Figure 3, Table S3). For those drugs tested by both disk diffusion and broth microdilution, the results correlated well between the two methodologies (Figure S2 and S3).

**Figure 3.**
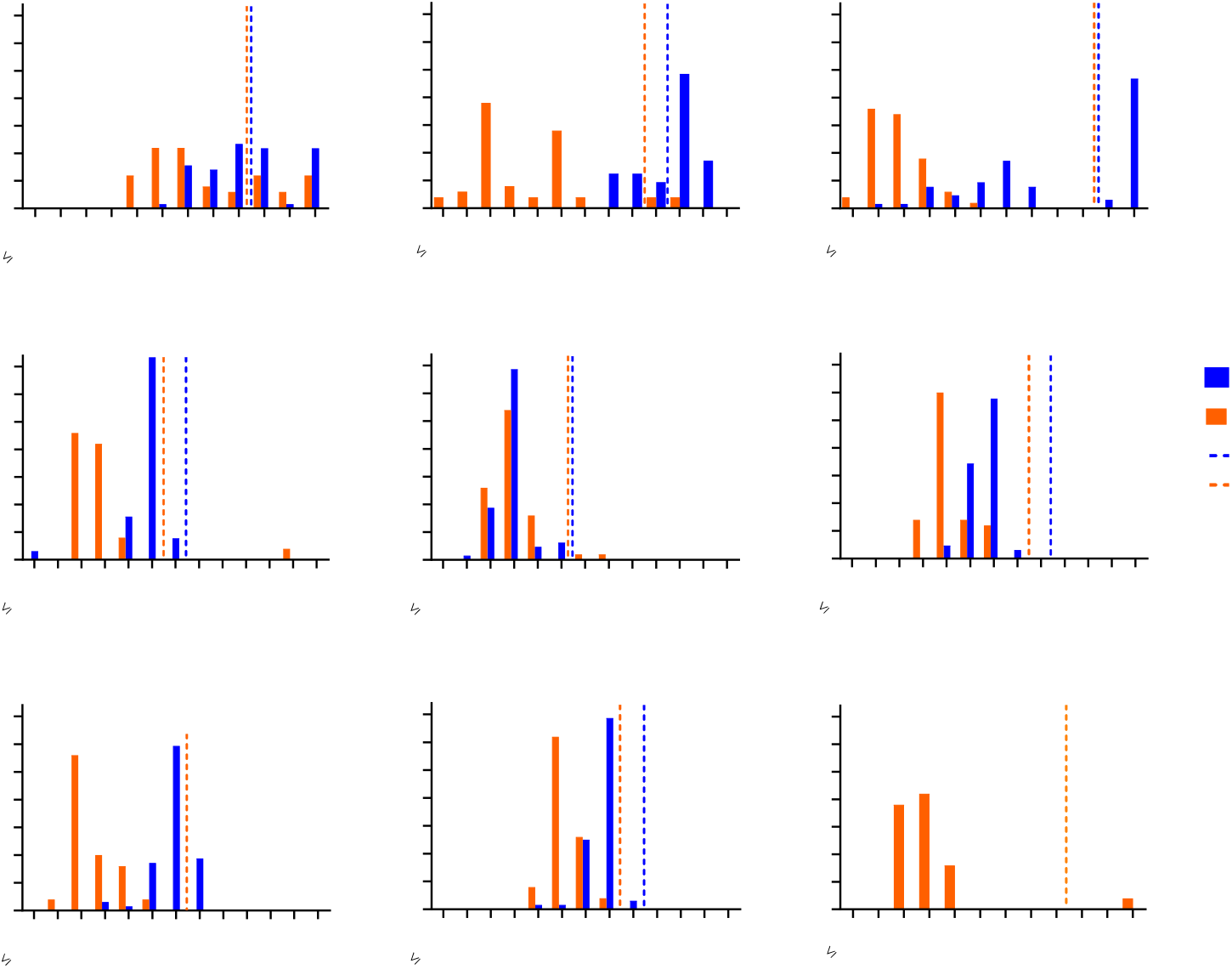
Minimal inhibitory concentration (MIC) distributions for respiratory *Enterobacterales* and *P. aeruginosa* isolates from ventilated COVID-19 patients. Vertical dashed lines indicate the EUCAST clinical resistant breakpoint values for piperacillin/tazobactam (TZP), cefepime (FEP), meropenem (MEM), gentamicin (GEN), tobramycin (TOB), amikacin (AMI) and trimethoprim/sulfamethoxazole (SXT) for *Enterobacterales* (orange) and *P. aeruginosa* (blue) isolates, respectively. In the case of plazomicin (PLZ), the vertical dashed line indicates the FDA-identified interpretative criteria. In the case of APR (apramycin), vertical dashed lines indicate the proposed ECOFF values [25].

Broth microdilution assays confirmed the results of the disk diffusion assay with regards to high aminoglycoside susceptibility of both *P. aeruginosa* and *Enterobacterales* and extended the aminoglycoside panel by including plazomicin (PLZ) and apramycin (APR). Both PLZ and APR showed full coverage of *Enterobacterales*. Apramycin additionally showed full coverage of *P. aeruginosa* (Figure 3, Table S3).

### Antibiotic resistance development in *P. aeruginosa* under antimicrobial selection pressure

Next, we analyzed bacterial isolates that were repetitively derived from individual patients to study changes to antimicrobial susceptibility in response to antibiotic therapy. A decrease in antimicrobial susceptibility over the course of treatment was particularly pronounced for *P. aeruginosa* (Figure S4). This prompted us to select a single *P. aeruginosa* infection for the analysis of phenotypic and genotypic changes in response to antibiotic therapy across eight consecutive isolates collected on day 9, 10, 16, 17, 18, 22, 29 and 37 (Figure 4).

**Figure 4.**
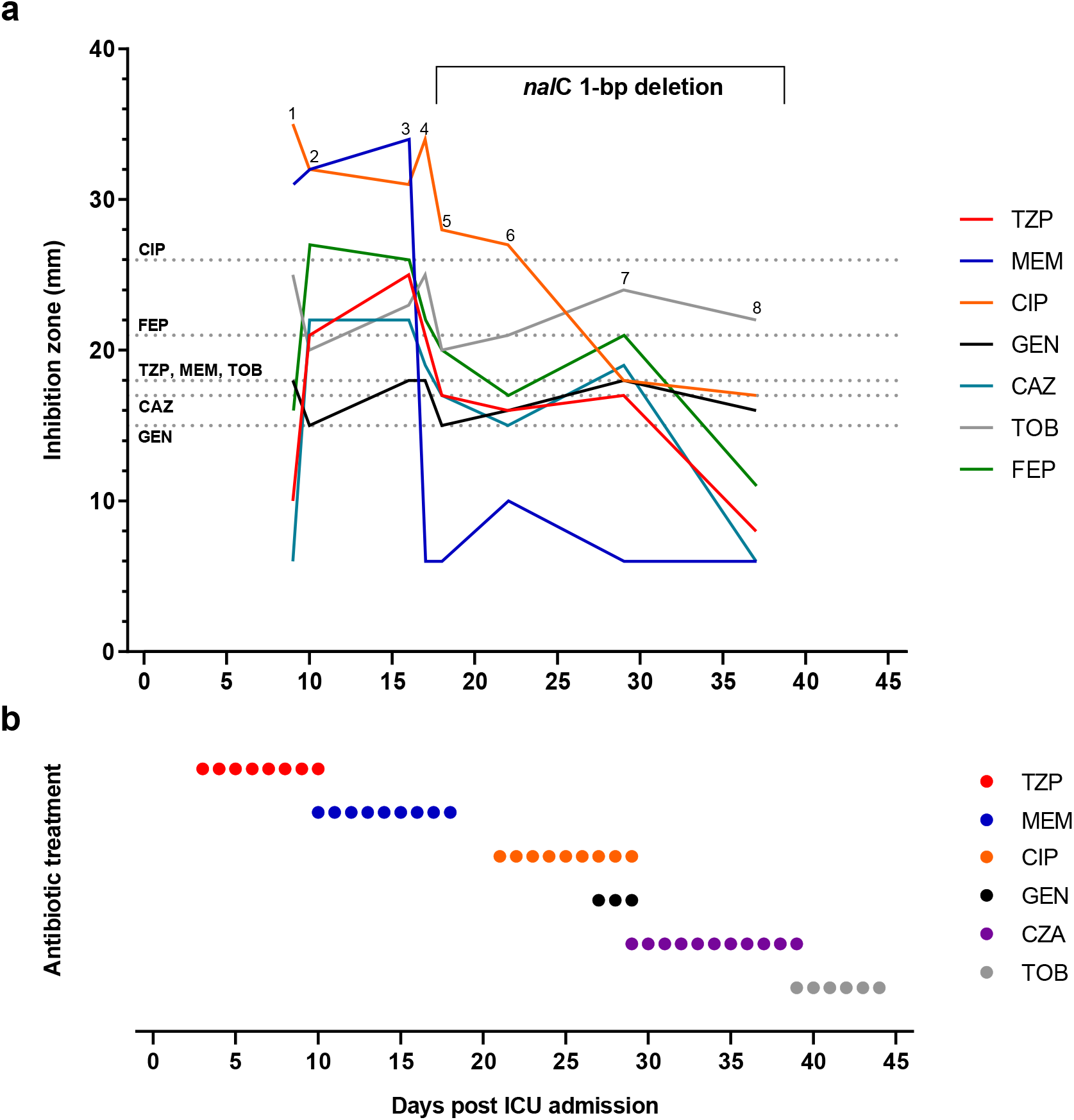
Antibiotic resistance development of *P. aeruginosa* in a single patient during antibiotic therapy. (a) The disc diffusion inhibition zones for piperacillin/tazobactam (TZP), meropenem (MEM), ciprofloxacin (CIP), ceftazidime (CAZ), cefepime (FEP), tobramycin (TOB) and gentamicin (GEN) are shown for eight consecutive isolates (labeled from 1-8), collected after 9, 10, 16, 17, 18, 22, 29 and 37 days of ICU admission. Horizontal dashed lines indicate the EUCAST clinical resistant breakpoint values for TZP (18mm), MEM (18mm), CIP (26mm), CAZ (17mm), FEP (21mm), TOB (18mm) and GEN (15mm), respectively. The bracket describes the resistance determinant detected in the *P. aeruginosa* isolates by whole-genome sequencing. (b) Antibiotic therapy of the patient. Piperacillin/tazobactam (TZP), meropenem (MEM), ciprofloxacin (CIP), gentamicin (GEN), ceftazidime/avibactam (CZA) and tobramycin (TOB).

The patient was initially admitted to the ICU of a regional hospital due to severe COVID-19 pneumonia. He had to be intubated and mechanically ventilated because of respiratory failure. After 2 days the patient was referred to the ICU of the University Hospital Zurich, a tertiary care hospital, because of pulmonary deterioration and worsening of inflammatory parameters and empiric antibiotic therapy for hospital-acquired pneumonia (HAP)/ventilator-associated pneumonia (VAP) was started with piperacillin/tazobactam. At this point, the patient had developed severe ARDS (oxygenation index 85 mmHg). Initial isolation of *Pseudomonas aeruginosa* from TBS occurred shortly after on day 4 after ICU admission (Figure S4, ID 05). After seven days of antibiotic therapy with piperacillin/tazobactam (TZP) resistance was not only detected against this broad-spectrum β-lactam antibiotic/β-lactamase inhibitor combination, but also against third and fourth generation cephalosporins (CAZ and FEP) (Figure 4). Consecutively, antibiotic therapy was switched to meropenem (MEM) and subsequent isolates (isolates 2 and 3) yet again showed susceptibility to TZP and cephalosporins (CAZ and FEP). After eight days of antibiotic therapy with MEM, repetitive isolates showed resistance to MEM (isolates 4 to 8) and eventually to TZP as well as extended-spectrum cephalosporins (CAZ and FEP), indicating a multidrug-resistant gram-negative (MDRGN) infection. In the meantime, the patient showed clinical improvement and had been successfully weaned from the respirator. Despite elevated inflammatory parameters the antimicrobial treatment was stopped for an antibiotics-free period. 2 days after stopping the antibiotic therapy, purulent endotracheal aspiration was obtained and therapy with ciprofloxacin (CIP) was initiated and continued for 9 days. Gentamicin was added because of pulmonary deterioration (Figure 4 and Figure S4, ID 05). Due to ongoing deterioration and after resistance to fluoroquinolones including CIP occurred, antibiotic therapy was switched to ceftazidime/avibactam (CZA) for a total of 11 days. Under antibiotic therapy with CZA the patient showed again clinical improvement but *P. aeruginosa* was still detectable throughout antibiotic treatment. Eventually the patient was successfully extubated on day 40 after initial ICU admission and intubation. Therapy with inhaled tobramycin (TOB) was started 2 days before extubation but was stopped again after a total of 6 days, as *P. aeruginosa* was still detectable and the patient showed no signs of acute infection, thus indicating ongoing colonization of the patient with MDR *P. aeruginosa*.

Finally, we performed whole-genome sequencing of the eight consecutive isolates in an attempt to detect possible resistance determinants that may explain the phenotypic progression (Excel file S1). Whole-genome analysis indicated that all eight isolates originated from the same clone indeed. The clone was characterized by two β-lactamases that commonly occur in *P. aeruginosa*: an OXA-50 like type and PDC-91. A 1-bp deletion causing a frameshift in the gene encoding the transcriptional regulator NalC of the *mexAB-oprM* multidrug efflux pump operon was found in isolates 4 till 8, but was absent in the first three isolates, providing a rationale for the drop in susceptibility from day 17 onwards (Figure 4).

## DISCUSSION

This study belongs to a group of early microbiological studies that report a detailed antimicrobial susceptibility profiling of Gram-negative bacterial superinfections in ventilated COVID-19 patients. The sample size is limited and has a strong geographic bias, but some important conclusions can nevertheless be drawn and will add to a growing number of similar studies that we expect from other geographic locations.

The etiology found here for late-onset VAP in COVID-19 patients resembles the diversity of Gram-negative pathogens typically found in nosocomial pneumonia [29]. Previous cohort studies have reported diverse etiology in COVID-19 confirmed cohorts, with *A. baumannii, K. pneumoniae, E. coli* and *P. aeruginosa* as predominant infecting agents [30–32].

Besides *P. aeruginosa* and *Enterobacterales, Burkholderia cenocepacia* were the third most common isolates encountered in this study. The latter is regarded a natural colonizer and an opportunistic pathogen in immune-compromised patients, with a naturally high intrinsic resistance to various antibiotics [33]. Trimethoprim-sulfamethoxazole (SXT) is used as a first-line option for the suppression/control of the infection. Since we only had a small number of *B. cenocepacia* isolates in our study, all of which were susceptible to SXT (Table S4), we decided to focus further analysis on only *P. aeruginosa* and *Enterobacterales*.

*P. aeruginosa* represents a notorious pathogen of nosocomial infections often characterized by MDR, especially in VAP and cystic fibrosis patients [34,35]. Treatment success is greatly hampered due to its intrinsic and adaptive resistance to nearly all available antipseudomonal agents. Important determinants driving resistance in *P. aeruginosa* are multidrug efflux pumps, alterations to its outer membrane porins, and the expression of β-lactamases [36]. For the *P. aeruginosa* isolates analyzed here, we found very high levels of drug resistance to TZP, FEP, CAZ and MEM.

For the *Enterobacterales* isolates we found a relatively high level of resistance to TZP, CRO, CAZ, and AMC. This may be explained by prior selection related to early antibiotic treatment in the course of COVID-19 infection, since AMC, CRO, and TZP are commonly used in Switzerland as first-line drugs for nosocomial bacterial pneumonia. Some of the species within the order of *Enterobacterales* are further characterized by a chromosomally encoded AmpC β-lactamase, which also contributes to decreased susceptibility to selected β-lactam antibiotics and in particular AMC [37]. The high resistance of *Enterobacterales* to amoxicillin/clavulanate (AMC, 72.0%) was not surprising considering intrinsic (chromosomal) AmpC genes of *E. cloacae, K. aerogenes*, and *C. freundii* [37]. *Enterobacterales* resistance to FEP, SXT, MEM and aminoglycosides was low or absent in comparison to the aforementioned antibiotics. Only a single *E. coli* isolate (Figure 2 and 3 and Figure S2), showed resistance to gentamicin and tobramycin, without prior exposure to aminoglycosides in the patient’s treatment regimen.

Our findings seem to be generally aligned with those of several other reports from countries that experienced a high incidence of severe COVID-19 cases, which proposed an increase in MDR bacterial infections in severely ill COVID-19 patients [32,38–41]. However, it is conceivable to assume that the species distribution and specific resistance patterns within individual studies may vary depending on the geographic and local ICU etiology of resistant isolates, study-specific patient recruitment and sample selection, and the treatment history of prior antibiotic exposure in regular VAP patients and COVID-19 patients.

There is no evidence to assume that the antimicrobial resistance rates in ventilated COVID-19 patients differ significantly from resistance rates from non-COVID-19 ventilated ICU patients and underlie mainly local ICU epidemiology. Surveillance of MDR infections in ventilated ICU patients during and after the COVID-19 pandemic is required to adequately monitor a putative increased incidence of MDR infections in this specific patient population.

In the present study, antibiotic resistance levels seemed to increase over the course of antibiotic treatment in the *P. aeruginosa* isolates (Figure 4 and Figure S4). Resistance to TZP but also to third- and fourth-generation cephalosporins (CAZ and FEP, respectively) occurred on day 7 of TZP therapy as reflected in isolate 1 (Figure 4). This may be attributable to the two β-lactamases (OXA-50 like type and PDC-91) which were identified by WGS in all eight isolates from that single infection. The OXA-50 like type enzyme is a class D β-lactamase, which has a reported narrow hydrolyzing spectrum including piperacillin but not meropenem [42]. PDC-91 is an inducible AmpC-like β-lactamase conferring resistance to broad-spectrum cephalosporins [43]. Surprisingly, we observed a completely susceptible phenotype in isolates 2 and 3. We hypothesize that these two isolates were colonizing a different niche in the airway tract, potentially with the formation of a biofilm, which could have limited antibiotic exposure and resulted in a sensitive phenotype in the disc diffusion assay.

The MexAB-OprM efflux pump is a clinically relevant efflux system in *P. aeruginosa* [37,44]. Transcription of the corresponding genes is under regulation of MexR, NalC and NalD. Overexpression of this efflux pump can be induced by mutations in *nalC*, leading to resistance against β-lactams (except imipenem) and quinolones but not aminoglycosides [45].

WGS analysis identified a 1-bp deletion in the suppressor gene *nalC* of isolates 4 – 8, which likely renders the corresponding protein non-functional and eventually results in over-expression of MexAB-OprM. Notably, at the same time we observed a gradual to massive decrease in TZP, CAZ, FEP, CIP and MEM susceptibility in the consecutive isolates (Figure 4), indicating the importance of the *nalC* mutation in developing the MDR phenotype of isolate 8. However, imipenem has been described as unaffected by the MexAB-OprM efflux pump [45]. Hence, the observed resistance to imipenem (Figure S4, ID 05) may be caused by a different resistance mechanism. In addition, expression levels of *mexAB-oprM* or of *oxa-485* and *pdc-91* have not been tested, therefore the precise correlation of each drug susceptibility to its resistance mechanism remains to be investigated. Nevertheless, the detected resistance determinants in combination with the antibiotic treatment history provide a rational explanation for the emergence of this MDR *P. aeruginosa* strain.

In the case of pneumonia, a main determinant of antibiotic efficacy is the drug concentration in the lung parenchyma tissue. Although intravenous aminoglycosides penetrate in lung parenchyma and bronchial secretions, measured lung tissue concentrations have been found to be relatively low for some aminoglycosides, because plasma concentrations are kept low in clinical care to avoid systemic toxicity [46]. While therapy with inhaled aminoglycosides are well described and tolerated in the treatment of cystic fibrosis, administration of nebulized aminoglycosides in acute pulmonary infections has largely remained an off-label domain and requires further evaluation [47]. Our results warrant further exploration of inhaled aminoglycosides as a critical component in the treatment of HAP/VAP with MDR GNB in COVID-19 patients. The drug candidate apramycin, currently in clinical development, has previously been suggested to have best-in-class coverage of drug-resistant GNB [25,48–50], which was confirmed for VAP bacterial isolates from COVID-19 superinfections here.

## CONCLUSION

In conclusion, resistance to first-line antibiotics was prevalent in bacterial isolates from ventilated COVID-19 patients in Switzerland during the first wave of COVID-19. It is conceivable that AMR plays a key role in those ventilated patients that contract a VAP despite empiric treatment, and contributes to morbidity and case fatality rates of patients that in addition to COVID-19 treatment are likely to receive multi-drug regimens of second-line or last-resort antibiotics to control the bacterial infection. Aminoglycosides have regained interest as potent broad-spectrum antibiotics in the face of β-lactam and specifically carbapenem resistance, and represent a treatment option less toxic than polymyxins. In the present study, Aminoglycosides were the most effective drug class *in-vitro* for the respiratory clinical isolates studied here, and the only drug class with full coverage of all *P. aeruginosa* isolates. However, given the relatively low pulmonary tissue penetration and the potential risk of adverse effects at higher dosing, their clinical utility including alternative ways of administration, such as inhalation, will need to be further evaluated in clinical trials for use in patients with resistance to second-line therapies.

## Data Availability

All relevant data has been made available in the manuscript and/or its supplementary information.

## LIST OF ABBREVIATIONS

AMI: Amikacin
AMC: Amoxicillin/clavulanate
APR: Apramycin
AST: Antimicrobial Susceptibility Testing
CIP: Ciprofloxacin
CLSI: Clinical and Laboratory Standards Institute
CRO: Ceftriaxone
CST: Colistin
CAZ: Ceftazidime
CZA: Ceftazidime/avibactam
ECOFF: Epidemiological Cutoff
EUCAST: European Committee on Antimicrobial Susceptibility Testing
FEP: Cefepime
GEN: Gentamicin
GNB: Gram-negative bacilli
MEM: Meropenem
MIC: Minimal Inhibitory Concentration
PLZ: Plazomicin
SXT: Trimethoprim/sulfamethoxazole
TOB: Tobramycin
TZP: Piperacillin/tazobactam

## DECLARATIONS

### Ethics approval and consent to participate

The study was approved by the Local Ethics Committee of the Canton of Zurich, Switzerland (Kantonale Ethikkommission Zurich BASEC ID 2020 – 00646). All necessary patient/participant consent has been obtained and the appropriate institutional forms have been archived.

### Consent for publication

All necessary patient/participant consent has been obtained and the appropriate institutional forms have been archived.

### Availability of data and material

Data generated or analyzed during this study are included in this published article and its supplementary information files. The genome sequencing files generated and analyzed during the current study are available from the corresponding author on reasonable request.

### Competing interests

Author SNH is a shareholder in Juvabis AG. All other authors declare no competing interests.

### Funding

This study was partially funded by Innosuisse (project no. 45893.1 INNO-LS). The work was supported by the Clinical Research Priority Program of the University of Zurich for the CRPP Precision medicine for bacterial infections; and by the University of Zurich, Institute of Medical Microbiology. SDB is supported by a Fellowship from the Promedica Foundation (1449/M). The funders had no role in study design, data collection and analysis, decision to publish, or preparation of the manuscript.

### Authors’ contributions

All authors contributed to the study conception and design. Material preparation, data collection and analysis were performed by Marina Gysin, Claudio Tirso Acevedo, Klara Haldimann, Elias Bodendoerfer, Frank Imkamp, Karl Bulut, Katja Becker, and Sven N. Hobbie. The first draft of the manuscript was written by Marina Gysin and all authors commented on previous versions of the manuscript. All authors read and approved the final manuscript.

## Acknowledgements

The authors would like to express their gratitude to Reinhard Zbinden, Annelies Zinkernagel and the diagnostic laboratory staff of the Institute of Medical Microbiology for their support in accessing the bacterial clinical isolates, in performing MALDI-TOF and disk diffusion assays, whole-genome sequencing, and in accessing anonymized clinical information. We would like to acknowledge special permit by the University of Zurich to conduct research at the university labs during the COVID-19 lockdown period.

## Notes

### Competing Interest Statement

SNH is a co-founder of Juvabis AG, a startup biotech company with an interest in aminoglycoside therapeutics. All other authors declare no competing interests.

### Funding Statement

This work was supported by an Innosuisse grant (project no. 45893.1 INNO-LS) and by the University of Zurich, Institute of Medical Microbiology. This work was also supported by the Clinical Research Priority Program of the University of Zurich for the CRPP Precision medicine for bacterial infections. The funders had no role in study design, data collection and analysis, decision to publish, or preparation of the manuscript. SDB is supported by a Fellowship from the Promedica Foundation (1449/M).

### Author Declarations

The study was approved by the Local Ethics Committee of the Canton of Zurich, Switzerland (Kantonale Ethikkommission Zurich BASEC ID 2020 - 00646)

