## Supplementary Information for "Antimicrobial susceptibility patterns of respiratory Gram-negative bacterial isolates from COVID-19 patients in Switzerland"

---

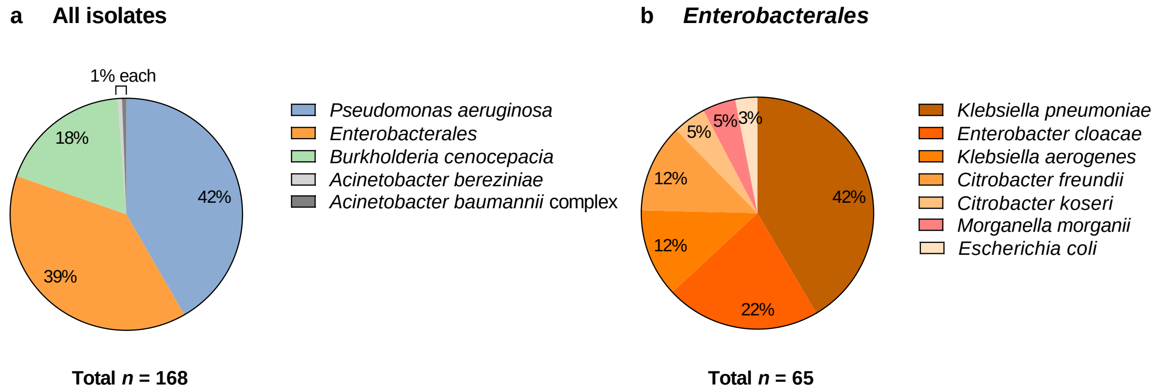

**Figure S1. MicrobiotaCOVID cohort species distribution of all identified Gram-negative isolates (a) and within *Enterobacterales* (b).**

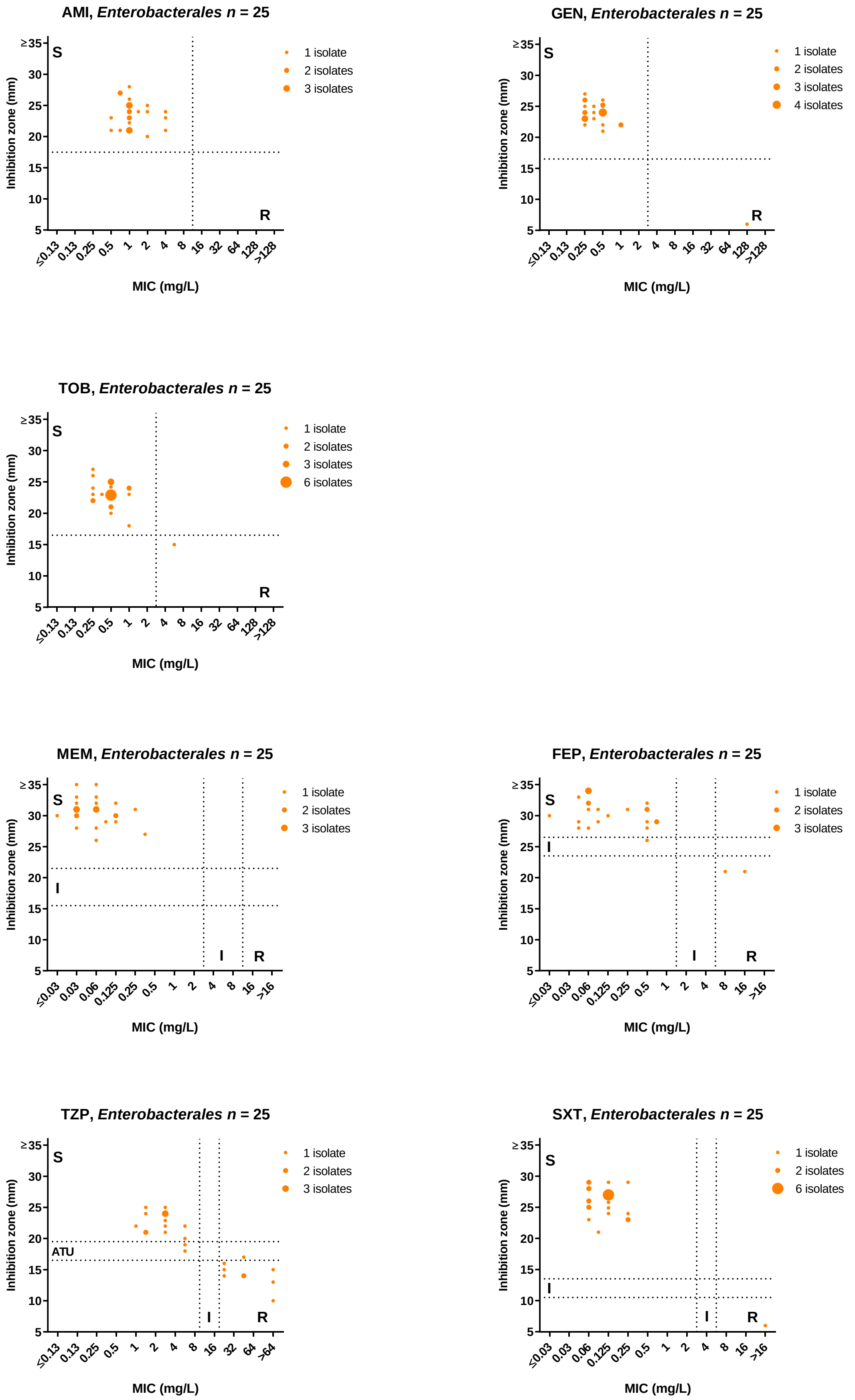

**Figure S2. Correlation plots of EUCAST disc diffusion and CLSI broth microdilution testing for *Enterobacterales*.** Amikacin (AMI), gentamicin (GEN), tobramycin (TOB), meropenem (MEM), cefepime (FEP), piperacillin/tazobactam (TZP) and trimethoprim/sulfamethoxazole (SXT).

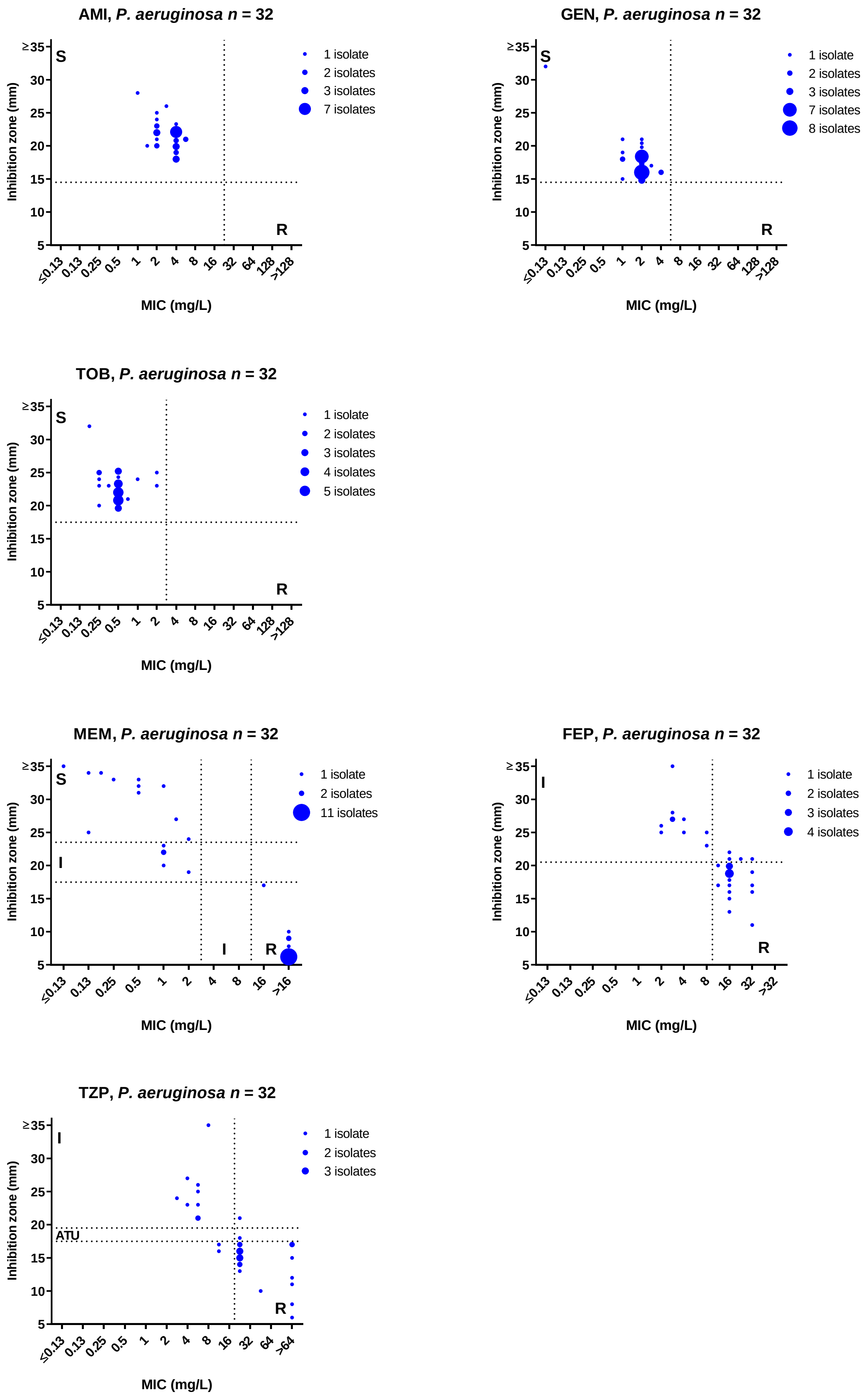

**Figure S3. Correlation plots of EUCAST disc diffusion and CLSI broth microdilution testing for *P. aeruginosa*.** Amikacin (AMI), gentamicin (GEN), tobramycin (TOB), meropenem (MEM), cefepime (FEP) and piperacillin/tazobactam (TZP).

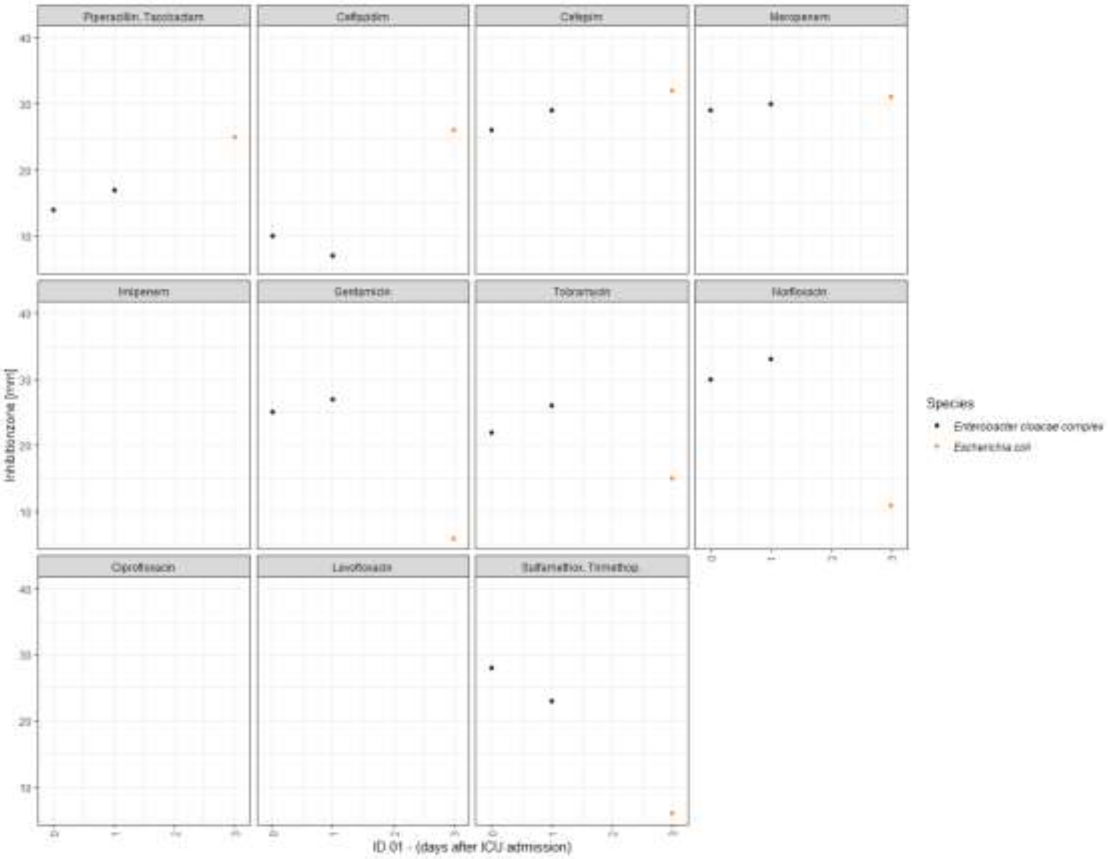

Piperacillin. Tazobactam
Meropenem
Imipenem
Tobramycin
Norfloxacin
Ciprofloxacin
Sulfamethox. Trimethop.
Inhibitionzone [mm]
Species
Enterobacter cloacae complex
ID 01 - (days after ICU admission)

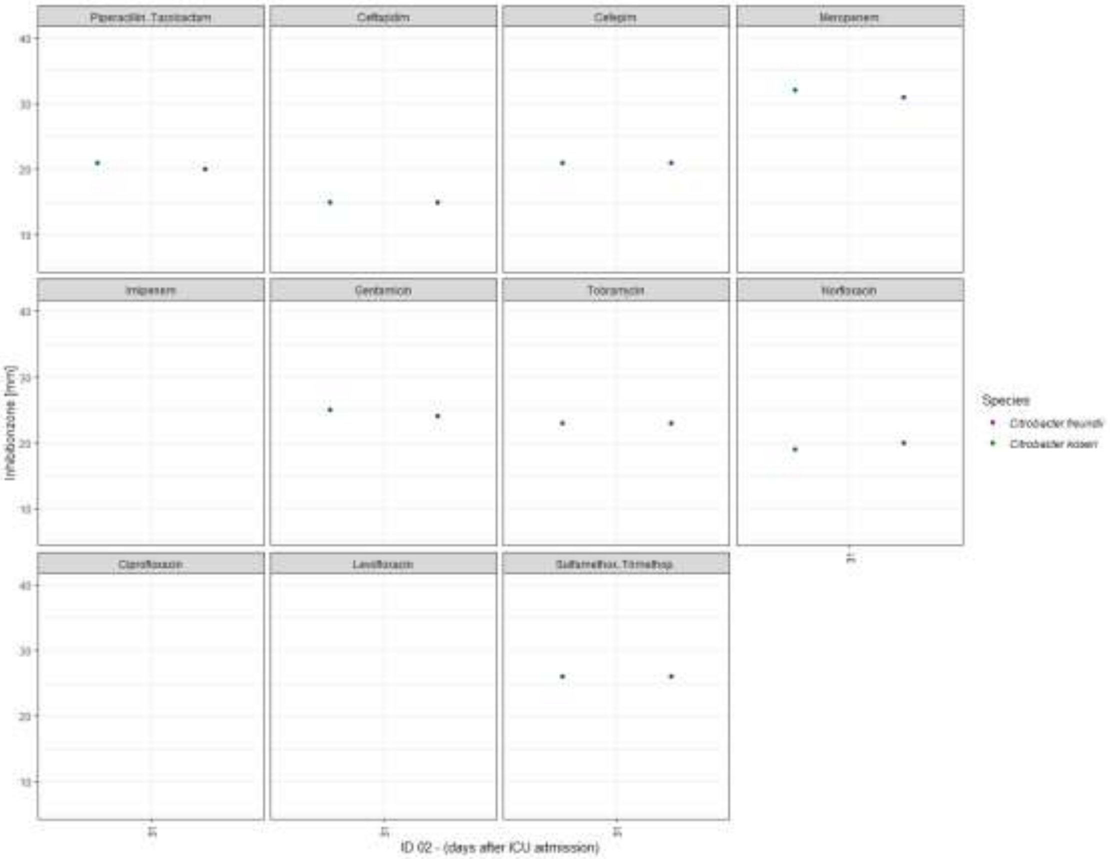

Piperacillin. Tazobactam
Meropenem
Imipenem
Gentamicin
Tobramycin
Norfloxacin
Ciprofloxacin
Sulfamethox. Trimethop.
Inhibitionzone [mm]
Species
ID 02 - (days after ICU admission)

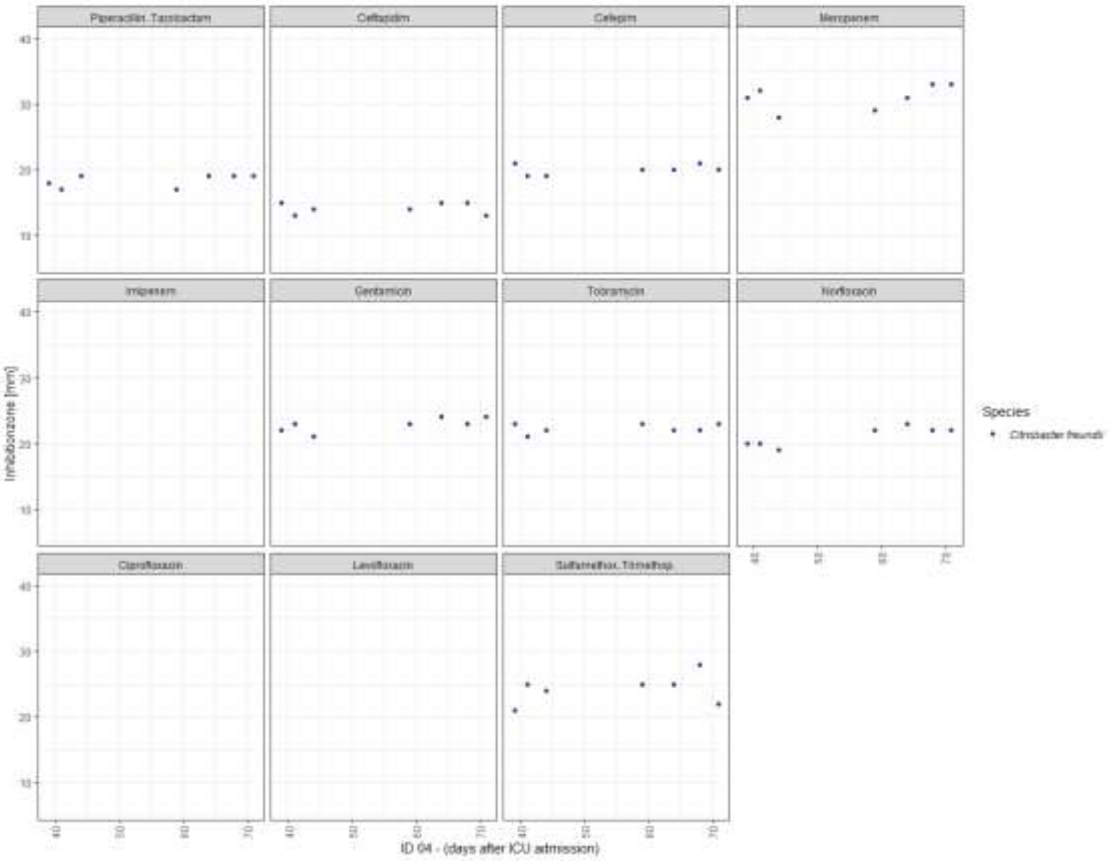

Piperacillin.Tazobactam
Ceftazidim
Cefepim
Meropenem
Imipenem
Gentamicin
Tobramycin
Norfloxacin
Ciprofloxacin
Levofloxacin
Sulfamethox.Trimethop.
Inhibitionzone [mm]
ID 04 - (days after ICU admission)
Species
Citrobacter freundii

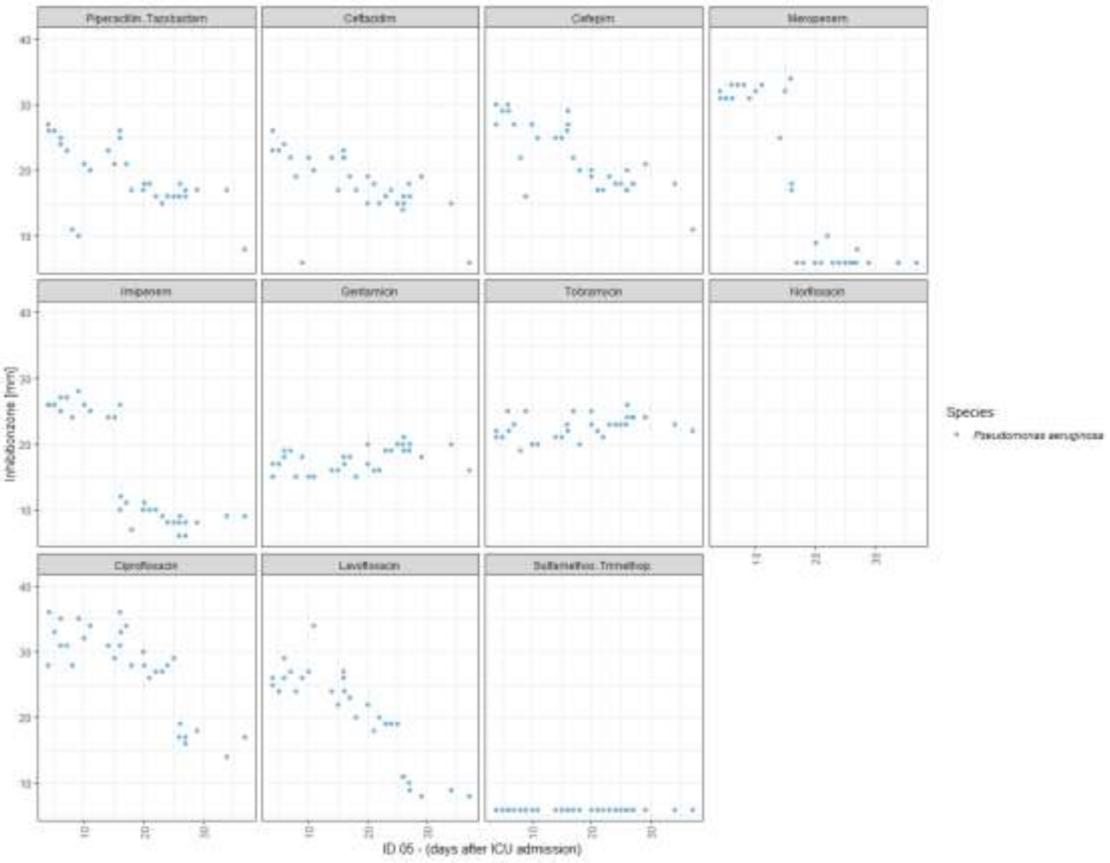

Piperacillin.Tazobactam
Ceftazidim
Cefepim
Meropenem
Imipenem
Gentamicin
Tobramycin
Norfloxacin
Ciprofloxacin
Levofloxacin
Sulfamethox.Trimethop.
Inhibitionzone [mm]
ID 05 - (days after ICU admission)
Species
Pseudomonas aeruginosa

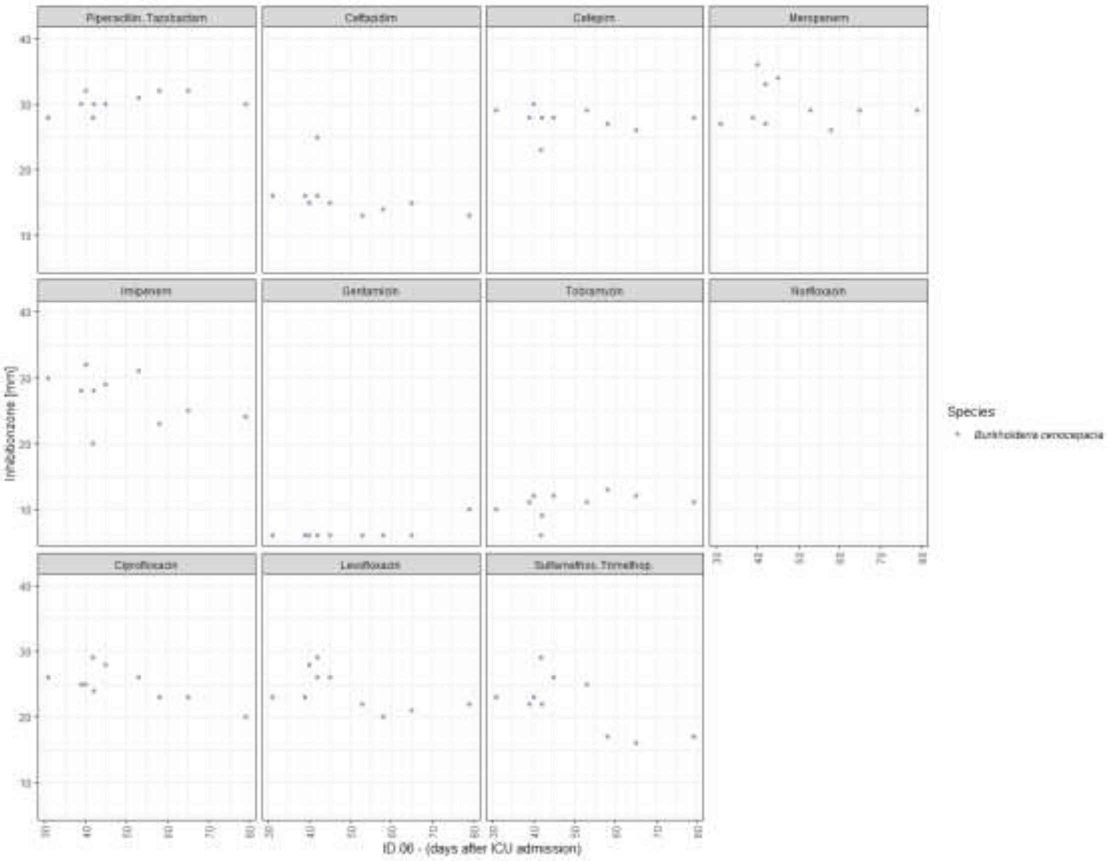
Piperacillin.Tazobactam
Ceftazidim
Cefepim
Meropenem
Imipenem
Gentamicin
Tobramycin
Norfloxacin
Ciprofloxacin
Levofloxacin
Sulfamethox.Trimethop.
Inhibitionzone [mm]
Species
Burkholderia cenocepacia
ID 06 - (days after ICU admission)

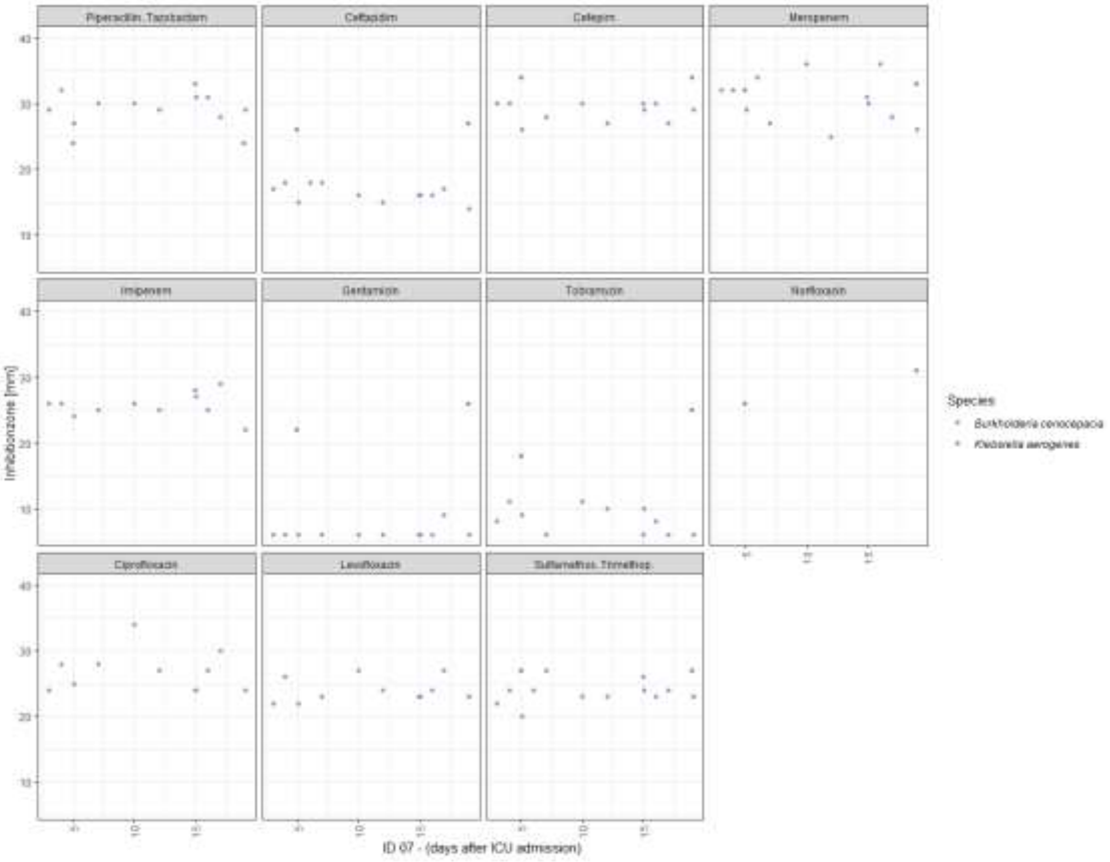
Piperacillin.Tazobactam
Ceftazidim
Cefepim
Meropenem
Imipenem
Gentamicin
Tobramycin
Norfloxacin
Ciprofloxacin
Levofloxacin
Sulfamethox.Trimethop.
Inhibitionzone [mm]
Species
Burkholderia cenocepacia
Klebsiella aerogenes
ID 07 - (days after ICU admission)

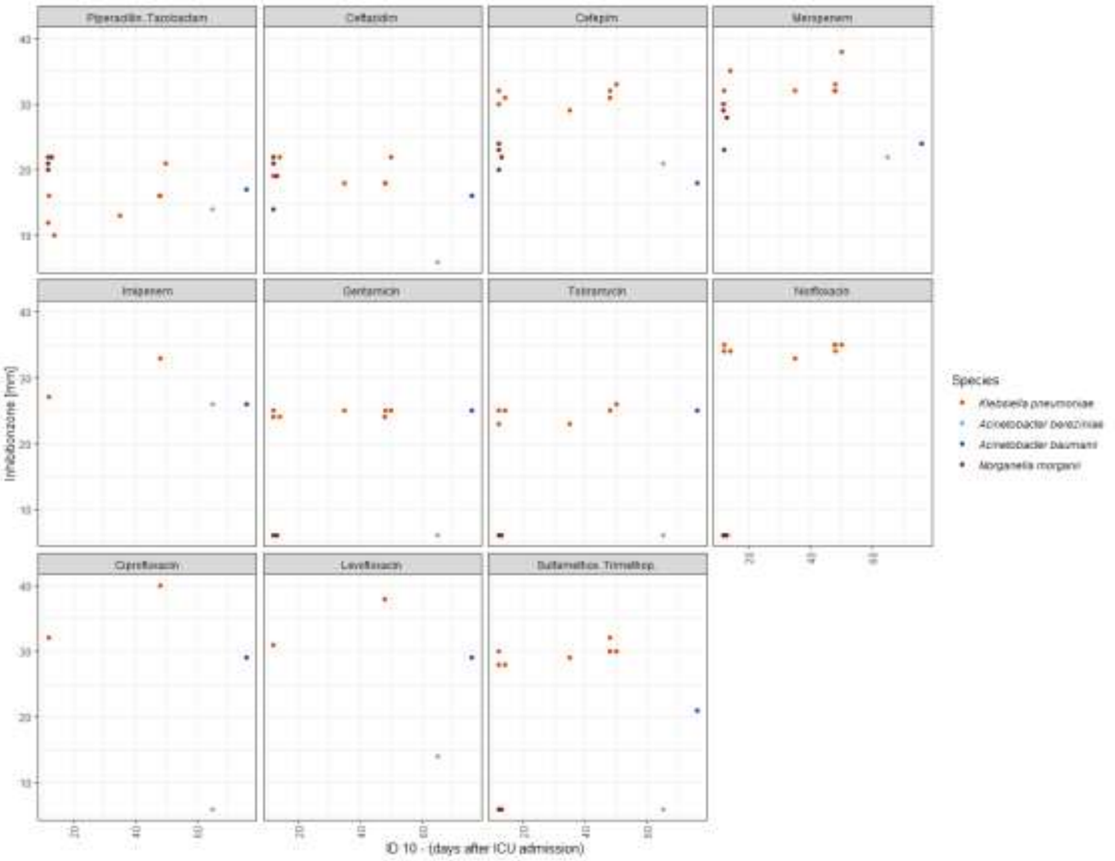
Piperacillin.Tazobactam
Ceftazidim
Cefepim
Meropenem
Imipenem
Gentamicin
Tobramycin
Norfloxacin
Ciprofloxacin
Levofloxacin
Sulfamethox..Trimethop.
Inhibitionzone [mm]
Species
Klebsiella pneumoniae
Acinetobacter baumanii
Morganella morganii
ID 10 - (days after ICU admission)

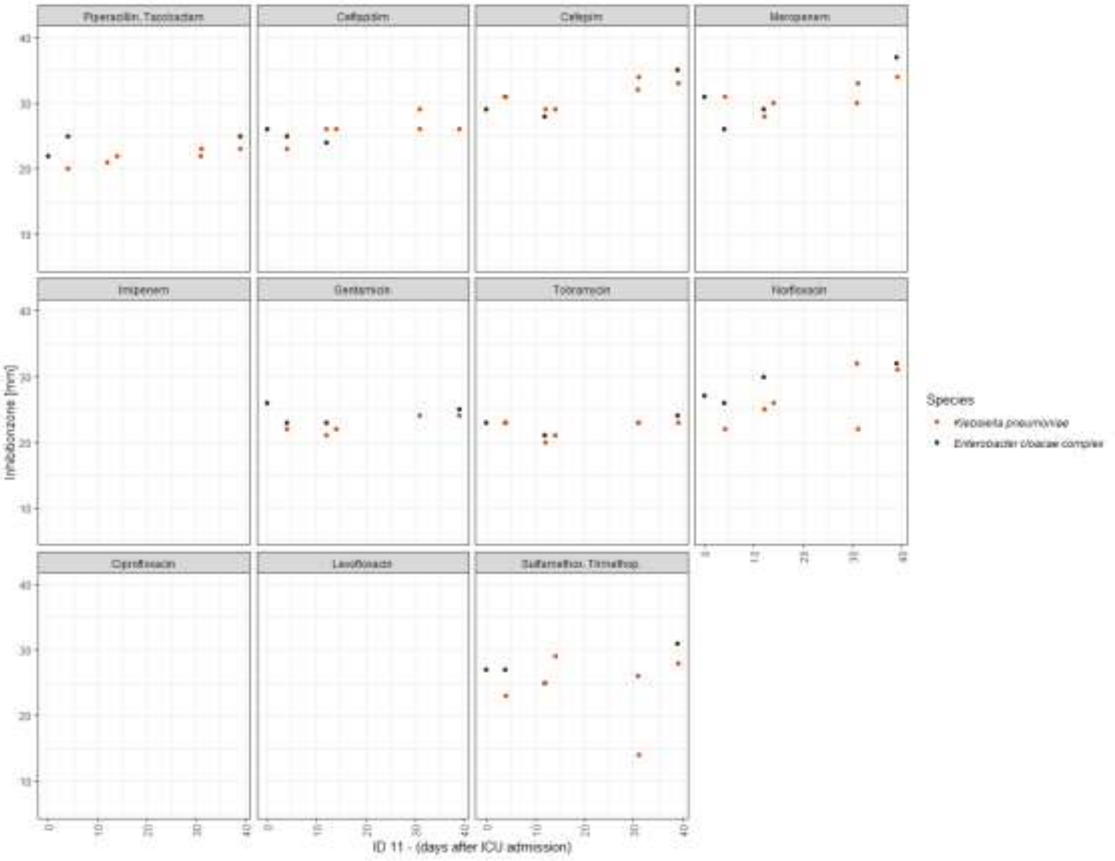
Piperacillin.Tazobactam
Ceftazidim
Cefepim
Meropenem
Imipenem
Gentamicin
Tobramycin
Norfloxacin
Ciprofloxacin
Levofloxacin
Sulfamethox..Trimethop.
Inhibitionzone [mm]
Species
Klebsiella pneumoniae
Enterobacter cloacae complex
ID 11 - (days after ICU admission)

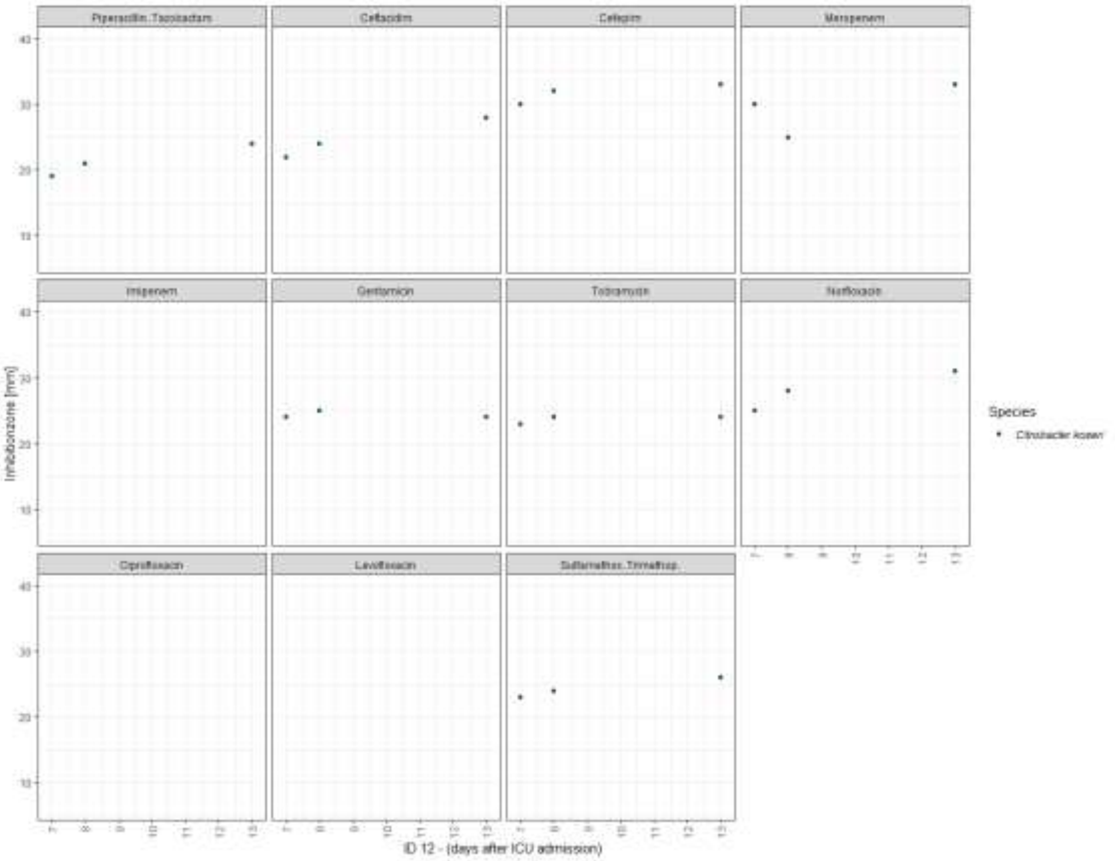
Piperacillin.Tazobactam
Ceftazidim
Cefepim
Meropenem
Imipenem
Gentamicin
Tobramycin
Norfloxacin
Ciprofloxacin
Levofloxacin
Sulfamethox.Trimethop.
Inhibitionzone [mm]
Species
Citrobacter koseri
ID 12 - (days after ICU admission)

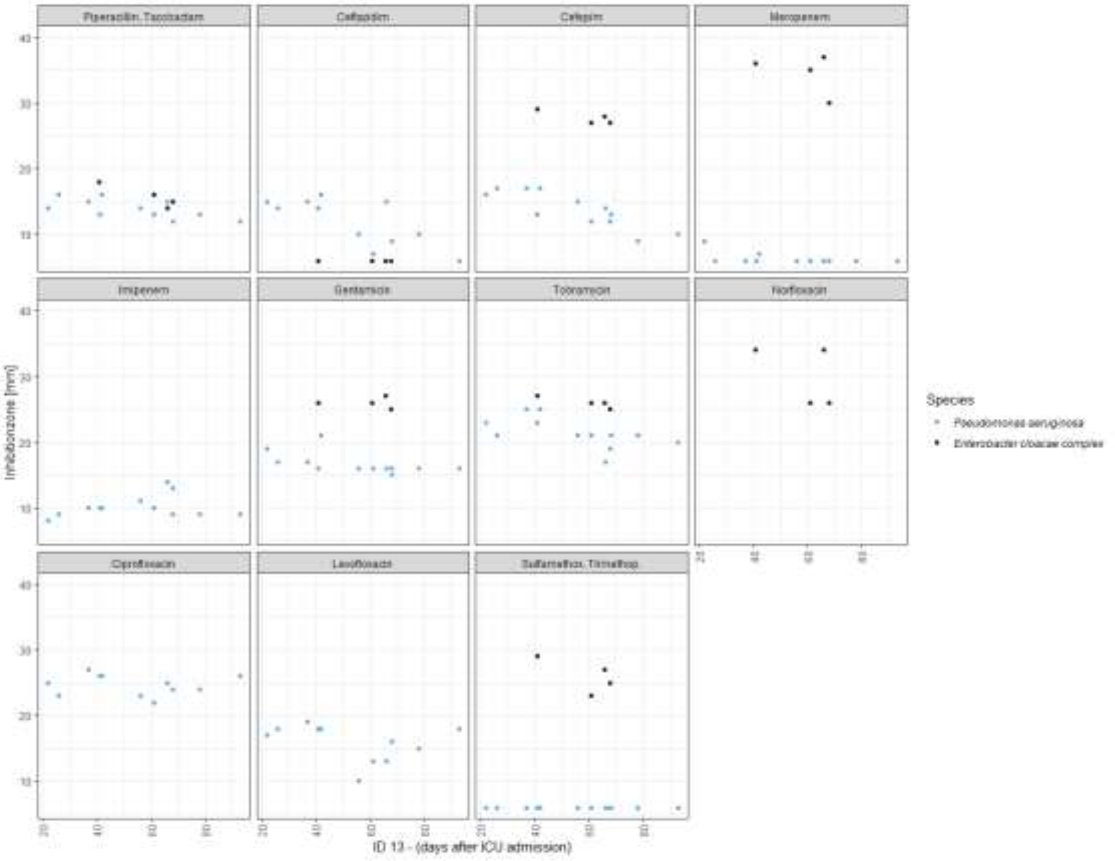
Piperacillin.Tazobactam
Ceftazidim
Cefepim
Meropenem
Imipenem
Gentamicin
Tobramycin
Norfloxacin
Ciprofloxacin
Levofloxacin
Sulfamethox.Trimethop.
Inhibitionzone [mm]
Species
Pseudomonas aeruginosa
Enterobacter cloacae complex
ID 13 - (days after ICU admission)

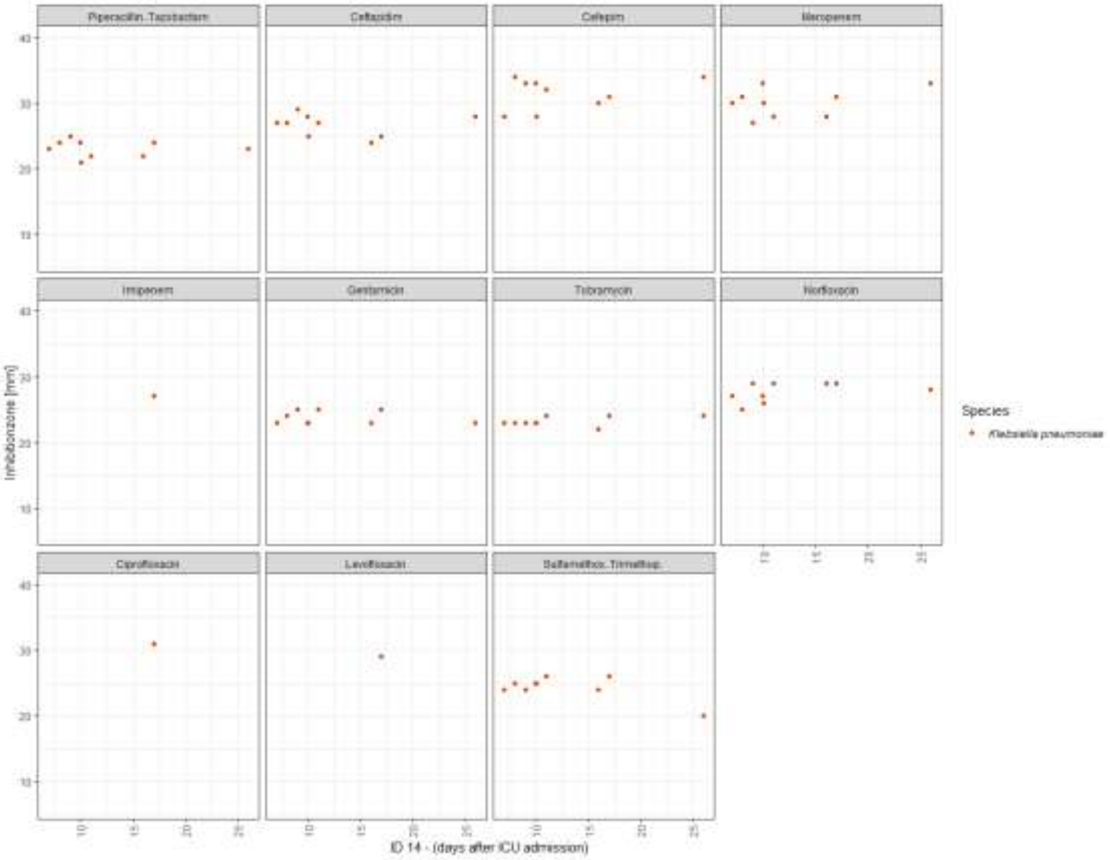

Piperacillin.Tazobactam
Ceftazidim
Cefepim
Meropenem
Imipenem
Gentamicin
Tobramycin
Norfloxacin
Ciprofloxacin
Levofloxacin
Sulfamethox. Trimethop.
Inhibitionzone [mm]
Species
Klebsiella pneumoniae
ID 14 - (days after ICU admission)

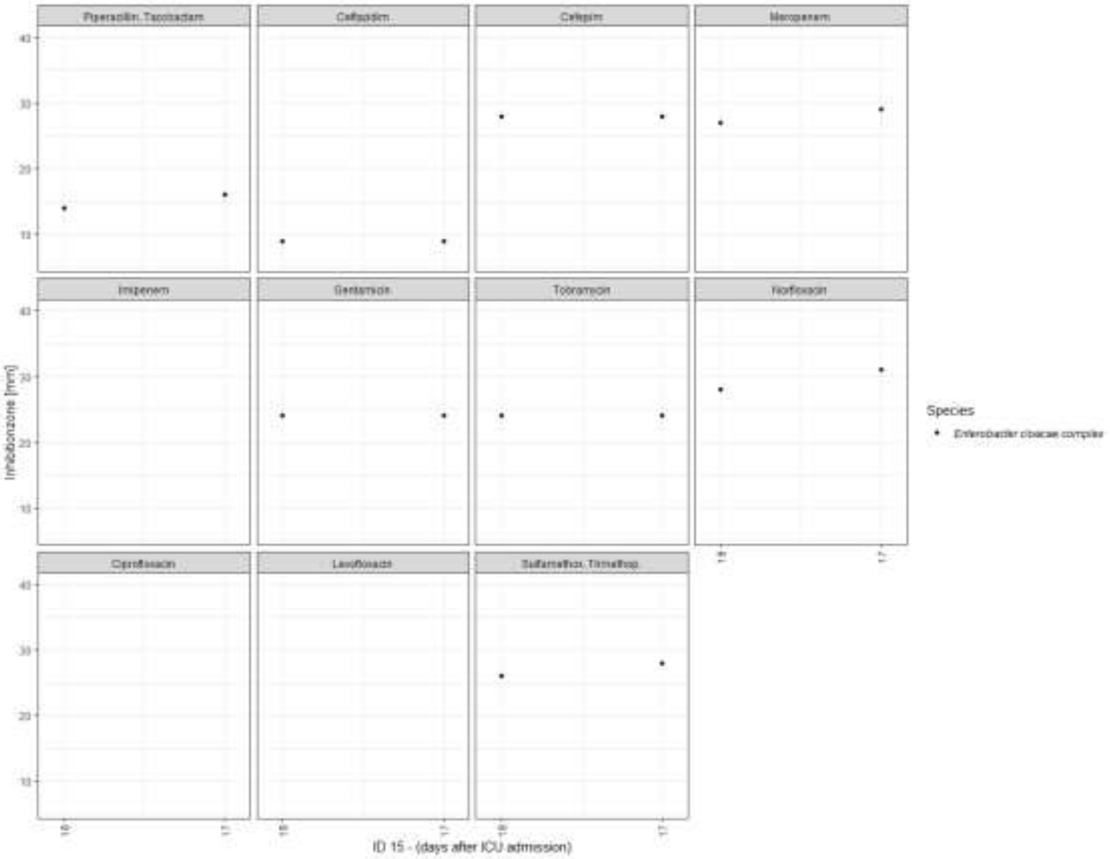

Piperacillin.Tazobactam
Ceftazidim
Cefepim
Meropenem
Imipenem
Gentamicin
Tobramycin
Norfloxacin
Ciprofloxacin
Levofloxacin
Sulfamethox. Trimethop.
Inhibitionzone [mm]
Species
Enterobacter cloacae complex
ID 15 - (days after ICU admission)

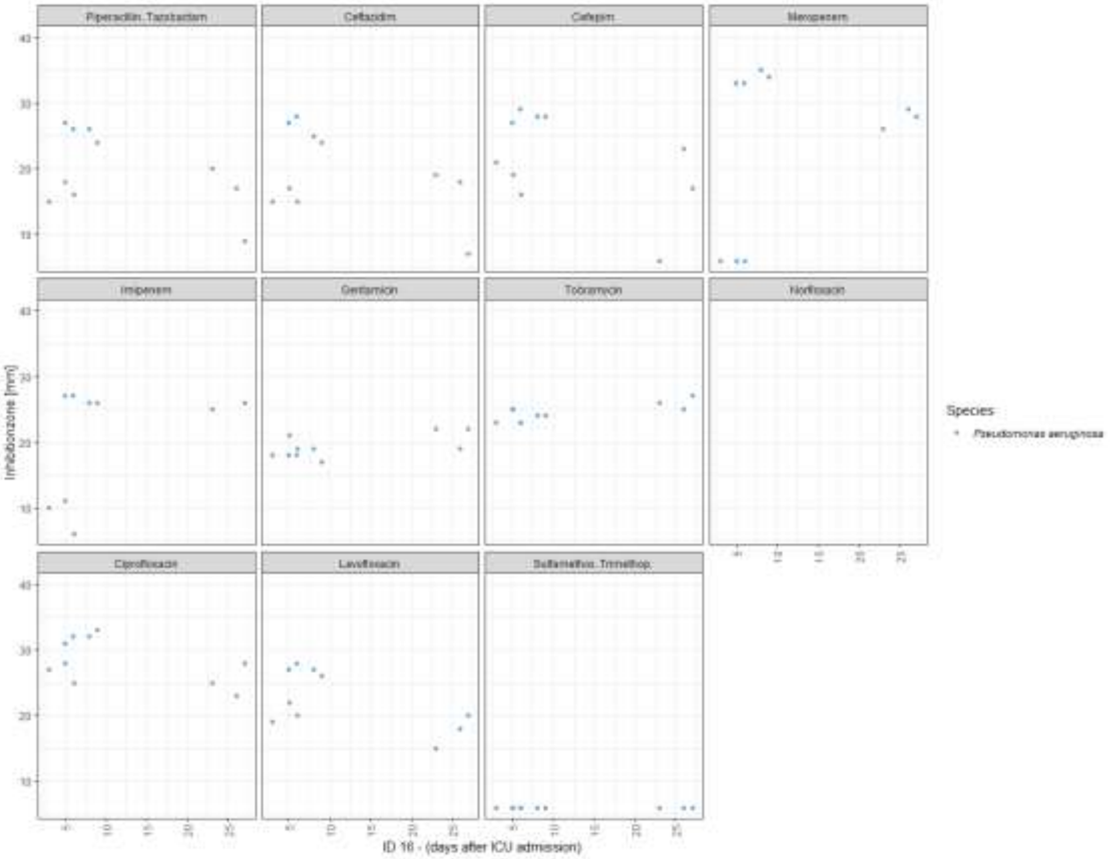
Piperacillin..Tazobactam
Ceftazidim
Cefepim
Meropenem
Imipenem
Gentamicin
Tobramycin
Norfloxacin
Ciprofloxacin
Levofloxacin
Sulfamethox..Trimethop.
Inhibitionzone [mm]
Species
Pseudomonas aeruginosa
ID 16 - (days after ICU admission)

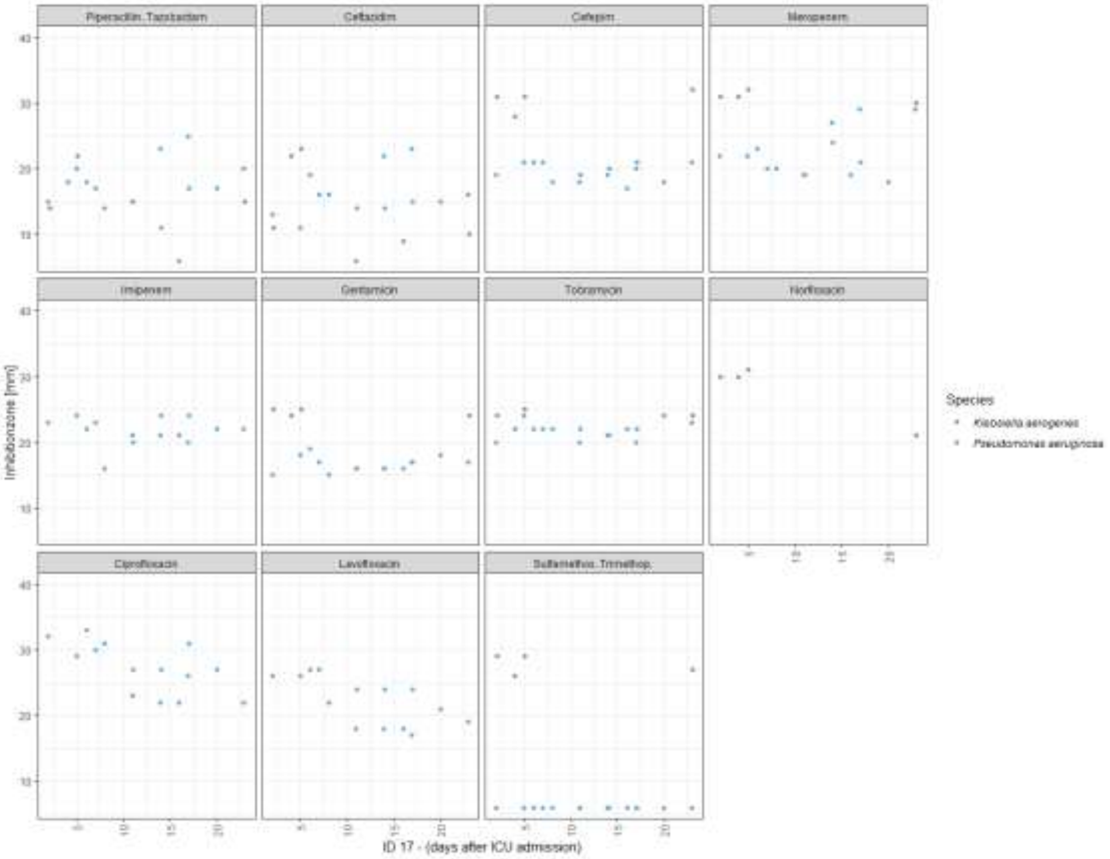
Piperacillin..Tazobactam
Ceftazidim
Cefepim
Meropenem
Imipenem
Gentamicin
Tobramycin
Norfloxacin
Ciprofloxacin
Levofloxacin
Sulfamethox..Trimethop.
Inhibitionzone [mm]
Species
Klebsiella aerogenes
Pseudomonas aeruginosa
ID 17 - (days after ICU admission)

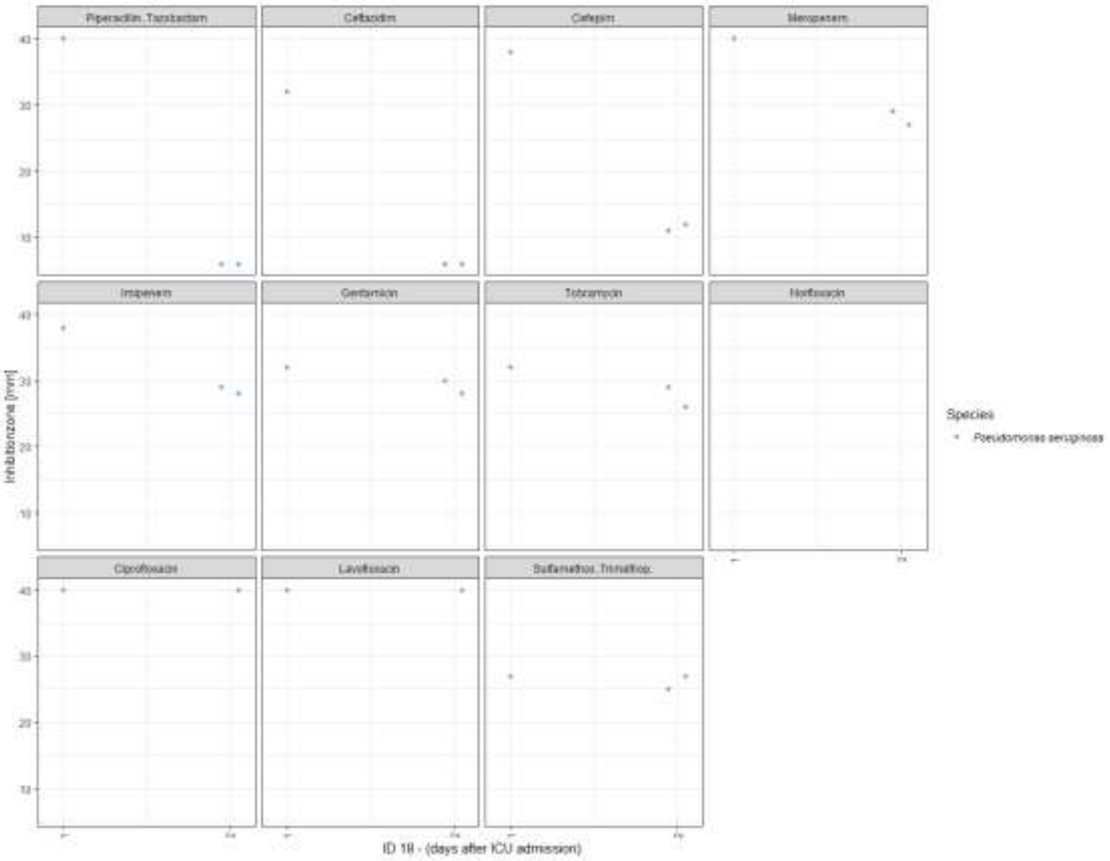

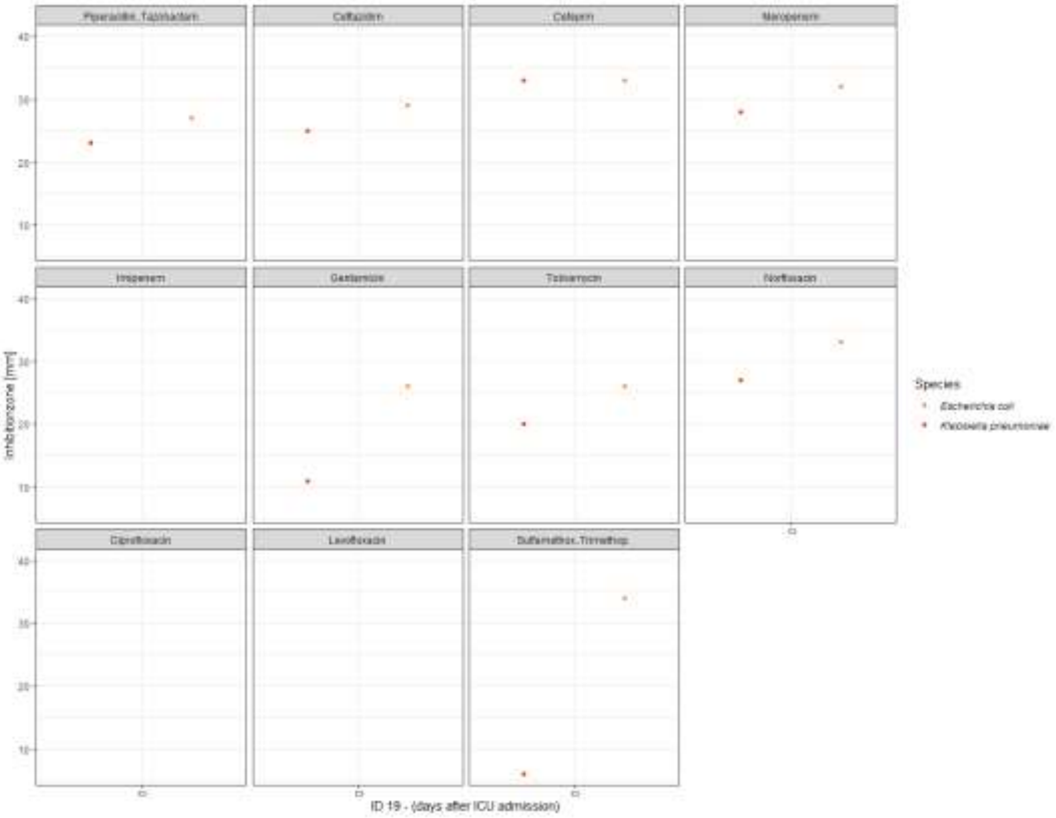

**Figure S4. EUCAST disc diffusion inhibition zones of repetitive microbiotaCOVID cohort isolates**. ID numbers refer to patient IDs (*n* = 16, only patients with repetitive isolates are shown).

**Table S1. Clinical characteristics of patients in this study (*n* = 17).**

| | ***n* = 17** |
|---|---|
| **Baseline characteristics** | |
| Age | 65 (56-59) |
| Male sex | 14 (82%) |
| Body mass index (kg/m2) | 27.8 (27.2-34.2) |
| **Clinical data at ICU admission** | |
| Sepsis-related organ failure assessment score (SOFA) | 9 (7-10) |
| Simplified acute physiology score (SAPS) II | 42 (28-51) |
| **Outcomes** | |
| Time of pulmonary superinfection detection after ICU admission (days) | 7 (3-16) |
| Duration of ventilation (days) | 37 (18-43) |
| Length of ICU stay (days) | 40 (24-51) |
| Patients died | 4 (24%) |
| **Antimicrobial therapy** | |
| Empiric antimicrobial therapy at ICU admission | 16 (94%) |
| Beta-lactam/beta-lactamase inhibitors (at any time) | 14 (82%) |
| Cephalosporins (at any time) | 15 (88%) |
| Carbapenems (at any time) | 12 (71%) |
| Aminoglycosides (at any time) | 8 (47%) |
| Quinolones (at any time) | 10 (59%) |

The data is presented as median (interquartile range (IQR)) or number and percentage (%).

**Table S2. Disc diffusion proportion of drug resistant and intermediate isolates**.

| | *Enterobacterales n* = 25 | | *P. aeruginosa n = 3*2 | |
|---|---|---|---|---|
| | **Disc diffusion** % resistant (*n*) | **Disc diffusion** % intermediate (*n*) | **Disc diffusion** % resistant (*n*) | **Disc diffusion** % intermediate (*n*) |
| FEP | 8.0% (2) | 4.0% (1) | 56.3% (18) | 43.8% (14) (r./i.) |
| TZP | 32.0% (8) | 12.0% (3) | 65.6% (21) | 34.4% (11) (r./i.) |
| SXT | 4.0% (1) | 0.0% (0) | No break point | No break point |
| MEM | 0.0% (0) | 0.0% (0) | 50.0% (16) | 15.6% (5) |
| AMI | 0.0% (0) | (n. ir.) | 0.0% | (n. ir.) |
| TOB | 4.0% (1) | (n. ir.) | 0.0% | (n. ir.) |
| GEN | 4.0% (1) | (n. ir.) | 0.0% | (n. ir.) |
| PLZ | No DD results | No DD results | No DD results | No DD results |
| APR | No DD results | No DD results | No DD results | No DD results |
| AMC | 72.0% (18) | 0.0% (0) | No DD results | No DD results |
| CRO | 32.0% (8) | 0.0% (0) | No DD results | No DD results |
| CAZ | 36.0% (9) | 0.0% (0) | 46.9% (15) | 53.1% (17) |
| CIP | No DD results | No DD results | 15.6% (5) | 84.4% (27) (r./i.) |

n.ir.: no intermediate disc diffusion range

r./i.: only resistant and intermediate disc diffusion range

cefepime (FEP), piperacillin/tazobactam (TZP), trimethoprim/sulfamethoxazole (SXT), meropenem (MEM), amikacin (AMI), tobramycin (TOB), gentamicin (GEN), plazomicin (PLZ), apramycin (APR), amoxicillin/clavulanate (AMC), ceftriaxone (CRO), ceftazidime (CAZ), ciprofloxacin (CIP)

**Table S3. MIC proportion of drug resistant and intermediate isolates.**

| | *Enterobacterales n* = 25 | | *P. aeruginosa n* = 32 | |
|---|---|---|---|---|
| | **MIC** % resistant (*n*) | **MIC** % intermediate (*n*) | **MIC** % resistant (*n*) | **MIC** % intermediate (*n*) |
| FEP | 8.0% (2) | 0.0% | 65.6% (21) | 34.4% (11) (r./i.) |
| TZP | 30.0% (7.5) | 6.0% (1.5) | 45.3% (14.5) | 54.7% (17.5) (r./i.) |
| SXT | 4.0% (1) | 0.0% (0) | No break point | No break point |
| MEM | 0.0% (0) | 0.0% (0) | 50.0% (16) | 0.0% (0) |
| AMI | 0.0% (0) | (n. ir.) | 0.0% (0) | (n. ir.) |
| TOB | 4.0% (1) | (n. ir.) | 0.0% (0) | (n. ir.) |
| GEN | 4.0% (1) | (n. ir.) | 0.0% (0) | (n. ir.) |
| PLZ | 0.0% (0) | 0.0% (0) | No break point | No break point |
| APR | 0.0% (0) | (n. ir.) | 0.0% (0) | (n. ir.) |

n.ir.: no intermediate MIC range

r./i.: only resistant and intermediate MIC range

cefepime (FEP), piperacillin/tazobactam (TZP), trimethoprim/sulfamethoxazole (SXT), meropenem (MEM), amikacin (AMI), tobramycin (TOB), gentamicin (GEN), plazomicin (PLZ), apramycin (APR)

**Table S4. MIC proportion of drug resistant and intermediate isolates.**

| | *B. cenocepacia n* = 12 | | | |
|---|---|---|---|---|
| | **Disc diffusion** % resistant (*n*) | **Disc diffusion** % intermediate (*n*) | **MIC** % resistant (*n*) | **MIC** % intermediate (*n*) |
| SXT | 0% (0) | 0% (0) | 0% (0) | (n.ir.) |
| MEM | 0% (0) | 0% (0) | 8.3% (1) | 25% (3) |

n.ir.: no intermediate MIC range

trimethoprim/sulfamethoxazole (SXT), meropenem (MEM)
